# Multi-trait genome-wide association study of opioid addiction: *OPRM1* and Beyond

**DOI:** 10.1101/2021.09.13.21263503

**Authors:** Nathan Gaddis, Ravi Mathur, Jesse Marks, Linran Zhou, Bryan Quach, Alex Waldrop, Orna Levran, Arpana Agrawal, Matthew Randesi, Miriam Adelson, Paul W. Jeffries, Emma C. Johnson, Nicholas G. Martin, Louisa Degenhardt, Grant W Montgomery, Leah Wetherill, Dongbing Lai, Kathleen Bucholz, Tatiana Foroud, Bernice Porjesz, Bradley Todd Webb, Richard C. Crist, Henry R. Kranzler, Hang Zhou, Gary Hulse, Dieter Wildenauer, Erin Kelty, John Attia, Elizabeth G. Holliday, Mark McEvoy, Rodney J. Scott, Sibylle G Schwab, Brion S. Maher, Richard Gruza, Mary-Jeanne Kreek, Elliot C. Nelson, Wade H. Berrettini, Joel Gelernter, Howard Edenberg, Laura Bierut, Dana B. Hancock, Eric O. Johnson

## Abstract

Opioid addiction (OA) has strong heritability, yet few genetic variant associations have been robustly identified. Only rs1799971, the A118G variant in *OPRM1*, has been identified as a genome-wide significant association with OA and independently replicated. We applied genomic structural equation modeling to conduct a GWAS of the new Genetics of Opioid Addiction Consortium (GENOA) data and published studies (Psychiatric Genomics Consortium, Million Veteran Program, and Partners Health), comprising 23,367 cases and effective sample size of 88,114 individuals of European ancestry. Genetic correlations among the various OA phenotypes were uniformly high (r_g_ > 0.9). We observed the strongest evidence to date for *OPRM1*: lead SNP rs9478500 (*p*=2.56×10^−9^). Gene-based analyses identified novel genome-wide significant associations with *PPP6C* and *FURIN*. Variants within these loci appear to be pleiotropic for addiction and related traits.

---

In 2020 the U.S. saw the highest 12-month count of opioid overdose deaths recorded, >70,000^1^, which represents a 40% increase since 2019, a >250% increase since 2000,^2^ and is 1.7 times the number of deaths caused by automobile crashes in 2020.^3^ Approximately 4% of the U.S. population aged 12 and older (10.1 million people) misused opioids in 2019, with 1.6 million people initiating new prescription opioid misuse.^4^ The most recent annual estimate of the total economic burden of prescription opioid abuse and dependence in the U.S. (2013) is over $78 billion,^5^ including Medicaid spending of more than $8 billion on opioid addiction (OA) treatment.^6^ By every metric, the opioid epidemic continues to be a tremendous burden, and the need to expand the medication-assisted treatment toolkit for OA through identification of new targets for drug development is clear.^7^

Animal model and human neuroimaging studies have established a strong, albeit partial, understanding of the neurocircuitry of addiction as heuristically characterized in the Koob and Volkow model. ^8^ The primary neurocircuitry elements involved (basal ganglia, extended amygdala, and prefrontal cortex) and their molecular connections to the cycle of addiction (intoxication, withdrawal, and preoccupation) are broadly understood. However, there is clear variability in the functioning of this neurocircuitry among individuals as evidenced by only 20–30% of people who use heroin becoming addicted ^9,10^ and only 8–12% of chronic pain patients prescribed opioids developing OA. ^11^

Genetics is a major contributor to individual variation in the risk of developing OA, with ∼60% of the population variability being attributable to genetic factors.^12,13^ This heritability estimate is comparable to other complex phenotypes, such as Alzheimer’s,^14^ age-related macular degeneration,^15^ and height,^16^ which have conclusively associated genetic variants. However, few robust genetic variant associations with OA have been identified.^17-20^

Eight genome-wide association studies (GWAS) of OA have been reported,^21-28^ in which the number of cases varied from 104 to 10,544 for ancestry specific analyses. Six of these GWAS identified genome-wide significant loci.^23,25-29^ However, only the largest analysis, which combined results of European ancestry (EA) cohorts from the US Veterans Affairs Million Veterans Program (MVP), the Study of Addiction: Genetics and Environment (SAGE), and Yale-Penn (YP) cohorts (10,544 cases and 72,163 controls), identified a genome-wide significant association that replicated in an independent sample (additional YP data: 508 cases and 206 controls). The variant identified is the long-studied rs1799971 (*OPRM1*-A118G), a functional coding variant (encoding Asn40Asp) in the mu opioid receptor gene (*OPRM1*): discovery *p*=1.51×10^−8^, replication *p*=0.049. The rs1799971-G protective association with OA was also extended at nominal significance to buprenorphine treatment status in the UK Biobank (240 cases and 360 901 controls; *p*=0.04).

To maximize discovery, we leveraged genomic structural equation modeling (gSEM)^30^ to combine new and existing GWAS with varied, but closely related, phenotypes for OA to enable the largest GWAS of OA to date (23,367 cases, 384,629 controls: effective sample size 88,114). We brought together novel results from the Genetics of Opioid Addiction Consortium (GENOA) with publicly available summary statistics from the MVP-SAGE-YP,^27^ the Psychiatric Genetics Consortium – Substance Use Disorder Group (PGC-SUD),^26^ and the Partners Health Group (PH).^28^ We examined SNP-based heritability and genetic correlation among the varied phenotypic definitions of OA across the contributing cohorts, including diagnostic and frequency of use-based cases and different types of controls: opioid exposed, unexposed, and population-based. We conducted a variant level gSEM analysis in the full complement of cohorts and a gene-based association test based on those results. gSEM accounts for the sample overlap among the GENOA, PGC-SUD, and MVP-SAGE-YP analyses, therefore increasing the available sample size compared to standard meta-analysis. Follow-up analyses included: (1) evaluation of genetic correlation with brain-related phenotypes; (2) estimation of predicted genetically driven differential expression in brain tissues; (3) colocalization of genetic association loci with cis-eQTLs; (4) evaluation of loci pleiotropy, and (5) druggability of nominated targets.

This study provides an unequivocal genome-wide significant association signal for the intron 1 locus in *OPRM1* and, through haplotype analysis, suggests that rs1799971 (A118G) may not be the driver of the locus’s association with OA. We further nominate two novel genome-wide significant gene-based associations with OA: *PPP6C* and *FURIN*. Both genes have been previously associated with phenotypes correlated with OA (e.g. *PPP6C* with cigarette smoking,^31,32^ alcohol consumption,^31^ and depressive symptoms;^33^ *FURIN* with schizophrenia,^34,35^ risk tolerance,^36^ and insomnia^36^). This study links these genes to predicted genetically driven differential expression in brain tissues by OA. Colocalization analysis supports a shared single variant between OA association and gene expression for *PPP6C* but provides less clear results for *OPRM1* and *FURIN*. Collectively, these results provide extended insight into the association of *OPRM1* with OA as well as novel genes associated with this phenotype.

## Results

### Different approaches to defining OA are highly genetically correlated

Our gSEM for OA brings together novel GWAS data from GENOA and summary statistics from all prior GWAS of OA that included more than 1,000 cases and 1,000 controls of European ancestry (EA).^26-28^ GENOA is a new consortium comprised of investigators who attend the National Institute on Drug Abuse Genetics and Epigenetics Cross-cutting Research Team Meetings and who have GWAS data on OA (Supplementary Table 1). In this study, OA refers to a broad meaning of addiction to opioids defined by multiple approaches to measuring the phenotype. The success of both the GENOA and gSEM analyses to maximize sample size and discovery then depends on similar heritability and high genetic correlations across the different measures of OA.

We focused on EA cohorts for the genetic correlation and gSEM analyses because these approaches, which allow us to maximize sample size by bridging phenotypes and accounting for cohort overlap, require linkage disequilibrium score regression (LDSC) results to model the genetic variance-covariance matrix. LDSC in turn depends on an ancestry-specific reference panel, which isn’t currently available for African Americans (AAs).

Among the 9 independent EA cohorts contributing to GENOA, OA was defined by Diagnostic and Statistical Manual criteria for opioid abuse or dependence (DSM-based; N=17,061) or by frequency of use (FOU) of illicit opioids (e.g., injecting heroin 10 or more times in the past 30 days; FOU-based; N=11,976; Supplementary Table 1). SNP-based heritability for both phenotypes was strong (DSM-based: *h*^*2*^=0.11, *SE*=0.03; FOU-based: *h*^*2*^=0.18, *SE*=0.04) and their genetic correlation robust (*r*_*g*_ =1.05, *SE*=0.16; SNP-based genetic correlations are not bound by 1.0).

Across the full set of GWAS results contributing to the gSEM GWAS (i.e., GENOA, MVP-SAGE-YP, PGC-SUD, and PH) there are additional OA definitions (MVP and PH used Electronic Health Record ICD-9 or ICD-10 codes for opioid use disorder [OUD]) and variation in type of controls used. GENOA cohorts used a combination of controls (opioid exposed, unexposed, and unknown exposure population controls). MVP and PH used opioid exposed controls, and the PGC-SUD results used here were based on unexposed controls. Regardless of the approach to defining OA or the type of controls, the LDSC genetic correlations across cohorts were very high (all pairwise *r*_*g*_ > 0.9, Supplementary Table 2). These heritabilities and genetic correlations show that the genetics contributing to OA are highly shared regardless of OA case definition or opioid exposure status of controls to which the cases are compared.

### GENOA GWAS identifies one European Ancestry specific OA association

Conducting ancestry specific and cross-ancestry meta-analyses of the GENOA cohorts (Supplementary Figures 1-6 and Supplementary Tables 3-5) yielded one genome-wide significant association locus on chromosome 4 among EAs (rs28386916-A, beta = 0.17, *p*=9.04×10^−9^). The rs28386916 variant was not associated with OA among AAs (beta = -0.025, *p*=0.51) and consequently was no longer significant in the EA+AA meta-analysis. rs28386916 is an intronic variant located within the long noncoding RNA ENST00000659878 and between the *SNCA* and *GPRIN3* genes (Supplementary Figure 7). Although, rs28386916-A is common (EUR MAF = 0.40; AFR MAF = 0.81) and was well imputed (imputation quality > 0.8 across cohorts), this variant is not available in the results from the independent MVP or PH GWAS.

### Genomic Structural Equation Model GWAS of Opioid Addiction identifies two other genome-wide significant loci in European Ancestry

A single common factor gSEM (Figure 1a) fit the GENOA, MVP-SAGE-YP, PGC-SUD, and PH summary statistics for OA well, with high Akaike information criterion and comparative fit index, and low standardized root mean squared root (SRMR) values (Figure 1a). Testing the association of 2.4 million variants available across all cohorts with the latent genetic factor (effective sample size N=88,114) identified two genome-wide significant loci (Figure 1b; Q-Q plot Supplementary Figure 8): one with 32 genome-wide significant variants (top variant rs9478500-C, beta = 0.136, *p*=2.56×10^−9^; Supplementary Table 6; forest plot Supplementary Figure 9a) on chromosome 6 and the other represented by a single variant on chromosome 16 (rs13333582-C, beta = -0.219, *p*=3.58×10^−8^; forest plot Supplementary Figure 10). The LDSC intercept for this model (Figure 1b), being approximately 1, indicates that these results are not due to uncontrolled inflation that one would expect from inadequately accounting for overlap in the cohorts contributing to some of the summary statistics used here.^30^

**Figure 1.**
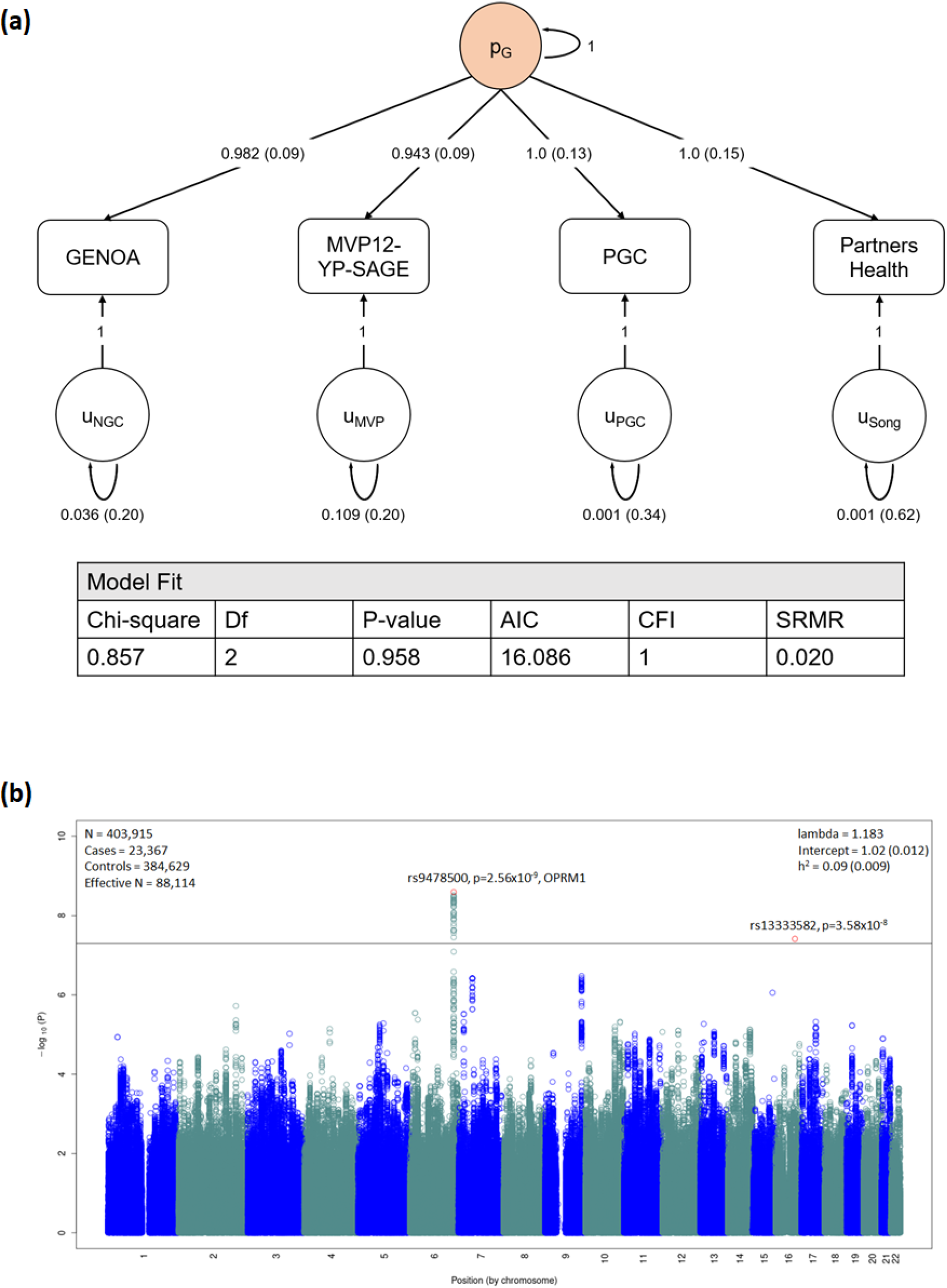
Genomic SEM model and Manhattan plot. (**a)** A common factor (p_g_) gSEM model (using GenomicSEM) is fit with summary statistics from GENOA, MVP12-YP-SAGE, PGC, and Partners Health cohorts. Standardized estimates and standard errors are shown for each free parameter. Model fit is shown by a non-significant chi-square test, high Akaike information criterion (AIC, higher is better) and comparative fit index (CFI) equal to exactly 1.0, and low standardized root mean squared root (SRMR) values (ideal < 0.05) (**b)** Manhattan plot for gSEM results with summary statistics from GWAS from each cohort. Bonferroni correction was used to correct for multiple comparisons; associations with P<2×10^−8^ (indicated by horizontal black bar) were considered to be genome-wide significant (top SNP highlighted in red).

The associated locus on chromosome 6 was centered in intron 1 of the mu-opioid receptor gene *OPRM1* (Supplementary Figure 11). The minor allele of the lead variant, rs9478500-C, was associated with increased risk of OA (beta = 0.136). All of the genome-wide significant variants were in high linkage disequilibrium (LD) with each other (r^2^ >0.88 and D’>0.93; Supplementary Table 7). The previously reported missense variant rs1799971 (*OPRM1*-A118G), which was genome-wide significant for OUD in MVP-SAGE-YP ^27^, was less statistically significant in our gSEM analysis (rs1799971-G, beta = -0.115, *p*=1.94×10^−6^; forest plot Supplementary Figure 9b). This variant has low r^2^ (<0.04), but perfect D’ (1.0), with the genome-wide significant variants observed here. In the MVP GWAS rs9478500-C was associated with OA, but with less statistical significance (MVP rs9478500-C, beta = 0.09, *p*=4.31×10^−5^). Prior candidate gene studies that examined *OPRM1* haplotypes with rs1799971 suggested that other variants may explain its equivocal association with OA.^37,38^ Raw data for haplotype analysis was available from a subset of cohorts contributing to the gSEM analysis (Figure 2). In this subset of cohorts, the single variant results for rs1799971 were weaker than in the gSEM (beta=-0.058, *p*=0.135) and slightly stronger for rs9478500 (beta=0.205, *p*=2.43×10^−9^). Comparison of the three haplotypes formed by rs1799971 and the genome-wide significant variants (Figure 2a) further weakened evidence for an association with OA being driven by rs1799971 (Figure 2b comparison 1: *p*=0.52, beta=-0.026, SE=0.0397) and strengthened evidence for an association with OA being driven by the effect of the non-rs1799971 variants (comparison 2: *p*=1.63×10^−10^, beta=0.2303, SE=0.036, Figure 2b).

**Figure 2.**
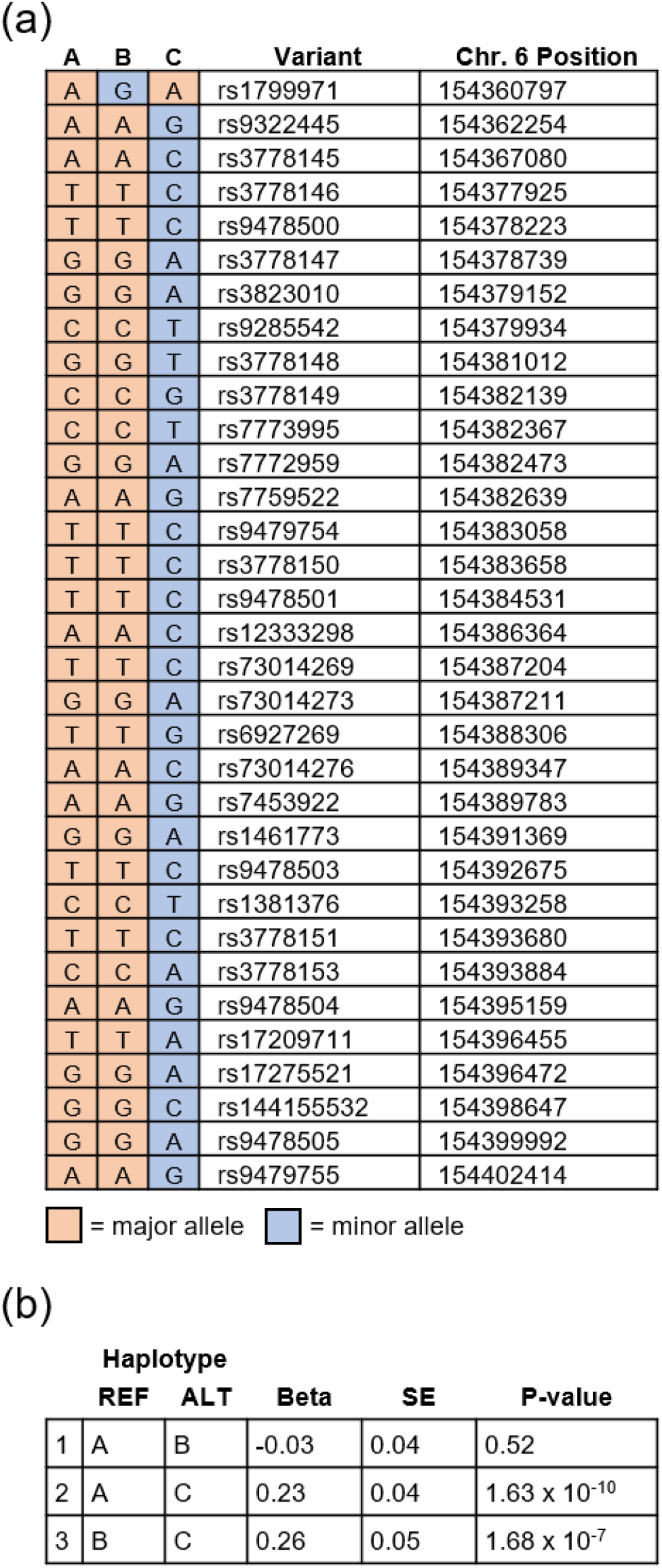
Association of major haplotypes for genome-wide significant *OPRM1* variants with OA. (a) The 3 major haplotypes for genome-wide significant *OPRM1* variants. Haplotype A is the predominant haplotype (frequency ∼0.69 among contributing cohorts) and consists of major alleles for all variants. Haplotype B (frequency ∼ 0.13 among contributing cohorts) consists of the minor allele for rs1799971 and the major allele for all other variants. Haplotype C (frequency ∼ 0.16 among contributing cohorts) consists of the major allele for rs1799971 and minor allele for all other variants. The cohorts for whom we had the raw data to conduct the haplotype analyses were: UHS, VIDUS, ODB, Yale-Penn, CATS and Kreek (Supplementary Table 1). (b) Association of *OPRM1* haplotypes with OA. Haplotype C is associated with increased risk of OA when compared to Haplotype A or Haplotype B, whereas Haplotype B does not have a significant impact on OA relative to Haplotype A. The single variant results using the cohorts contributing to the haplotype analyses were: rs1799971 beta=-0.058, *p*=0.135; rs9478500 beta=0.205, *p*=2.43×10^−9^.

The genome-wide significant rs13333582 variant on chromosome 16 is intergenic (Supplementary Figure 9): rs13333582-C minor allele (frequency = 0.04, imputation quality > 0.8 for all cohorts) being associated with decreased risk of OA (beta = -0.22). However, the variants with which rs13333582 has strong LD showed weak evidence for association with OA (e.g., rs921982 r^2^ = 0.87, p=2.25×10^−3^).

### OA is genetically correlated with 21 other brain-related traits

We used LDSC to estimate the genetic correlation between OA (gSEM results) and 37 brain related traits (Figure 3, Supplementary Table 8); of these, 21 were significantly correlated with OA at the Bonferroni corrected threshold of *p*<1.35×10^−3^. We observed high positive genetic correlations for OA with cannabis use disorder and alcohol dependence, as well as modest positive correlations across smoking traits and psychiatric disorders. Expected inverse genetic correlations were also evident for age of initiation of cigarette smoking and cognitive/educational traits. There were no genetic correlations between OA and brain volume traits.

**Figure 3.**
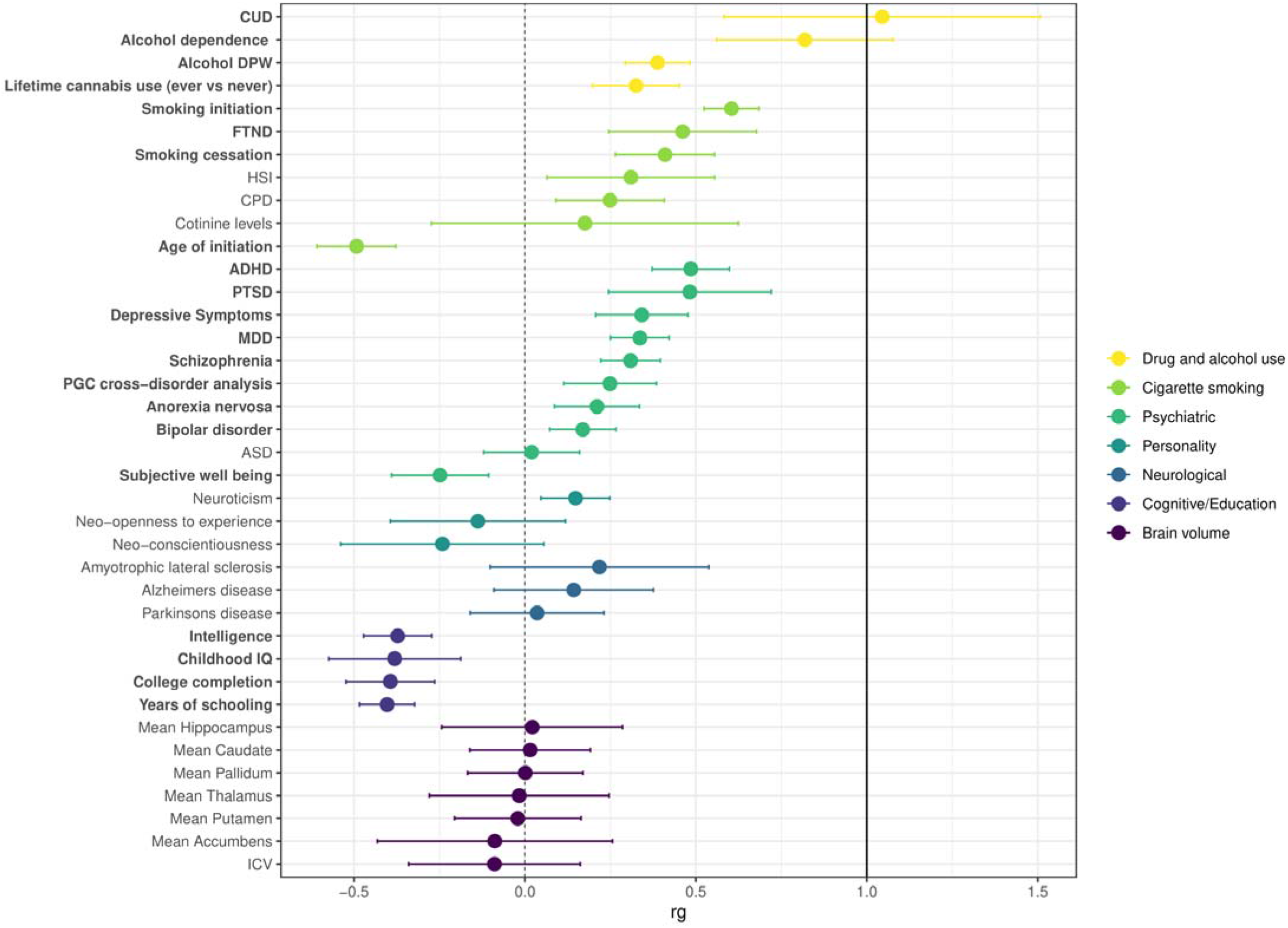
Genetic correlations of opioid addiction (OA) with 38 other brain-related phenotypes. Correlations were calculated using linkage disequilibrium (LD) score regression with the gSEM OA GWAS meta-analysis results, compared with results made available via LD Hub or study investigators (see Supplementary Table 20 for original references). Phenotypes were grouped by disease/trait or measurement category, as indicated by different colorings. Dots indicate the mean values for genetic correlation (*r*_g_); error bars show the 95% confidence intervals; the dashed vertical black line corresponds to *r*_g_□=□0 (no correlation with OA), and the solid vertical black line corresponds to *r*_g_□=□1.0 (complete correlation with OA). Phenotypes with significant correlation with OA are bolded (1 degree of freedom Chi-square test; Bonferroni adjusted *p*-value <0.05 after accounting for 38 independent tests). Exact *p*-values are provided in Supplementary Table 8).

### Gene-based MAGMA GWAS of gSEM summary statistics for OA corroborates *OPRM1* and identifies novel genes

To enhance statistical power for discovery, we analyzed the summary statistics from the gSEM GWAS at the gene level using MAGMA ^39^. In addition to *OPRM1*, we observed two novel genes associated with OA that surpassed Bonferroni correction for the 15,977 genes tested (*p*<3.13×10^−6^; Figure 4; Q-Q plot Supplementary Figure 13; full results Supplementary Table 9).

**Figure 4.**
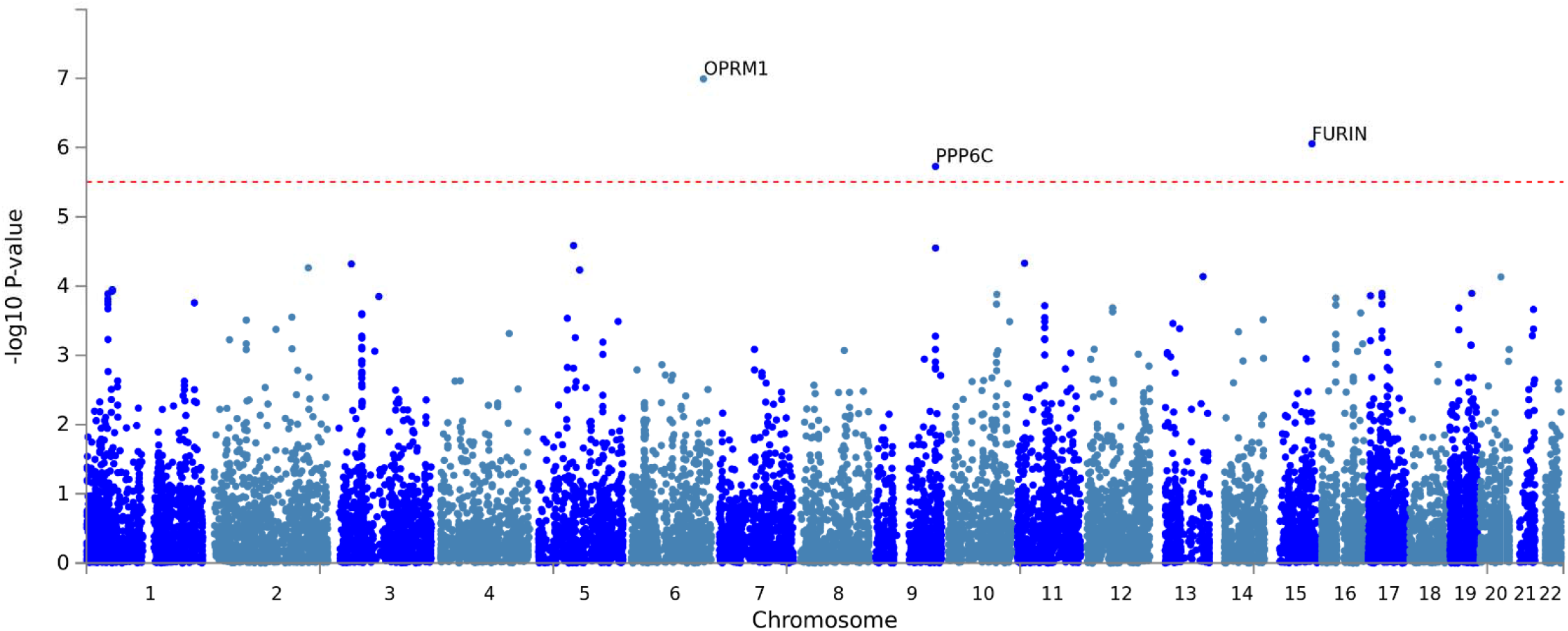
Gene-level Manhattan Plot. GWAS results were summarized at the gene-level using MAGMA. Bonferroni correction was used to correct for multiple comparisons; associations with P<3×10^−6^ (indicated by horizontal red dotted line) were considered to be genome-wide significant.

The gene-level association between the Protein Phosphatase 6 Catalytic subunit gene (*PPP6C*) and OA was based on 57 variants and corresponds to the variant-level peak that approached genome-wide significance on chromosome 9 (Figure 1b). This peak encompassed *PPP6C* but variants in high LD and with moderate p-values for association with OA (*p*=3.37×10^−7^ to 2.14×10^−5^) extended across three genes: *PPP6C*, the Suppressor of Cancer Cell Invasion gene [*SCAI*], and the Rab9 Effector Protein with Kelch Motifs gene [*RABEPK*] (Supplementary Figure 14). However, *SCAI* and *RABEPK* associations with OA did not surpass Bonferroni correction (*SCAI p*=0.0016; *RABEPK p*=2.82×10^−5^)

The gene-level association for the Furin Paired Basic Amino Acid Cleaving Enzyme gene (*FURIN*) and OA was based on a single variant (rs17514846-A, beta = -0.08, *p*=8.82×10^−7^). Other *FURIN* variants were excluded from the gSEM GWAS, and thereby the MAGMA analysis, due to the gSEM method’s requirement that variants be present in every contributing cohort. By running a standard logistic regression meta-analysis of *FURIN* variants across the subset of GWAS cohorts without overlapping participants (GENOA, MVP, and PH), we were able to retain additional variants excluded from the gSEM analysis, and identified 3 additional variants in strong LD with rs17514846 (r^2^>0.64, D’=1.0); all four variants were associated with OA (Supplementary Figure 15; Supplementary Table 10), the weakest association being for rs17514846 (*p*=1.67×10^−6^) and the strongest being for rs11372849, which was genome-wide significant (rs11372849-TC, beta = -0.074, *p*=4.11×10^−8^; forest plot Supplementary Figure 16).

### Predicted genetically driven gene expression in brain tissue expands neurobiologically relevant evidence for OA-associated genes

To estimate genetically driven differential gene expression in human brain tissues associated with OA, we applied S-PrediXcan ^40^ using GTEx version 8 eQTL gene models (http://predictdb.org/) with the gSEM GWAS summary statistics as input. Fourteen gene-tissue combinations surpassed correction for the total number of gene models and brain tissues (156,215 tests) with an FDR < 0.05 (Table 1; all results presented in Supplementary Table 11). Predicted genetically driven *OPRM1* expression was significantly associated with OA in cerebellum. Only four brain tissues had gene models for *OPRM1* (cerebellum, cerebellar hemisphere, hypothalamus, and nucleus accumbens; Supplementary Table 11). In contrast, 12 brain tissues had gene models for *PPP6C*; of these, *PPP6C* was predicted to be differentially expressed in nine tissues. Nearby *SCAI* was the only other gene to show statistically significant genetically driven expression associated with OA, doing so across four brain tissues. *FURIN* was nominally associated (best *p*=9.67×10^−5^ in hippocampus) but did not surpass the FDR<0.05 threshold. *RABEPK* was not predicted to be differentially expressed by OA (best p=0.055 in caudate).

**Table 1.** Fourteen gene-brain region combinations exhibiting predicted genetically driven differential gene expression in human brain regions associated with OA (across tissue FDR < 0.05) in analysis of gSEM GWAS summary statistics with S-PrediXcan analysis using GTEx version 8 eQTL gene models.

| Gene | Tissue | Across Tissue FDR |
| --- | --- | --- |
| *OPRM1* | Cerebellum | 0.009 |
| *SCAI* | Cerebellum | 0.009 |
| *SCAI* | Frontal cortex | 0.009 |
| *SCAI* | Hippocampus | 0.009 |
| *PPP6C* | Hippocampus | 0.009 |
| *PPP6C* | Anterior cingulate cortex | 0.01 |
| *PPP6C* | Cerebellar hemisphere | 0.01 |
| *PPP6C* | Putamen basal ganglia | 0.01 |
| *PPP6C* | Caudate basal ganglia | 0.01 |
| *SCAI* | Cortex | 0.01 |
| *PPP6C* | Cortex | 0.01 |
| *PPP6C* | Frontal cortex | 0.01 |
| *PPP6C* | Hypothalamus | 0.01 |
| *PPP6C* | Nucleus accumbens basal ganglia | 0.01 |

### Some OA associations colocalize with genetically driven gene expression

To estimate the likelihood that the genetic loci associated with OA share a causal variant with the expression quantitative trait loci (eQTLs) for our nominated genes (*OPRM1, PPP6C*, and *FURIN*), we applied coloc ^41^ to our gSEM GWAS results and the GTEx eQTL results for these genes. Because the variants underlying the genome-wide significant association for *PPP6C* physically extend into *SCAI* and *RABEPK* (Supplementary Figure 14), we included these genes in the analysis. We evaluated colocalization for these genes across the superset of 10 brain tissues which showed genetically driving differential expression for at least one gene in the S-PrediXcan analysis (Supplementary Table 11). *OPRM1* is expressed at relatively low levels in the GTEx brain tissues (Supplementary Figure 17a). Only six of 10 brain tissues showed variant associations with *OPRM1* expression in GTEx and could be included in the coloc analysis. The posterior probabilities for four tissues of the six tissues tested for *OPRM1* favored the hypothesis that only a genetic association with OA at this locus is present (Figure 5 H2, Supplementary Table 12). However, in the cerebellum, where *OPRM1* is most highly expressed and for which S-PrediXcan predicted differential expression by OA, the greatest posterior probabilities favored hypotheses for both the OA-associated locus and *cis*-eQTL traits being associated, but with different causal variants (H3) or a shared single causal variant (H4). Among the three genes at the *PPP6C*-centered locus, *PPP6C* shows the highest levels of gene expression in brain tissues (Supplementary Figure 17b-d) and the greatest support for colocalization of OA-associated variants with *cis*-eQTLs for *PPP6C* (Figure 5, Supplementary Table 12). In contrast, the analysis for *RABEPK* uniformly indicated that the OA-associated variants do not colocalize with the *RABEPK cis*-eQTLs. For *FURIN*, the null hypothesis of neither trait being associated in this region has the highest posterior probability across all tested brain tissues (Figure 5, Supplementary Table 12), which is consistent with (1) a single variant (rs17514846) driving the genome-wide significant gene-based association with OA, (2) no significant evidence for differential gene expression in the S-PrediXcan analyses, (3) limited evidence for this variant as an eQTL in brain tissues (Supplementary Table 13).

**Figure 5.**
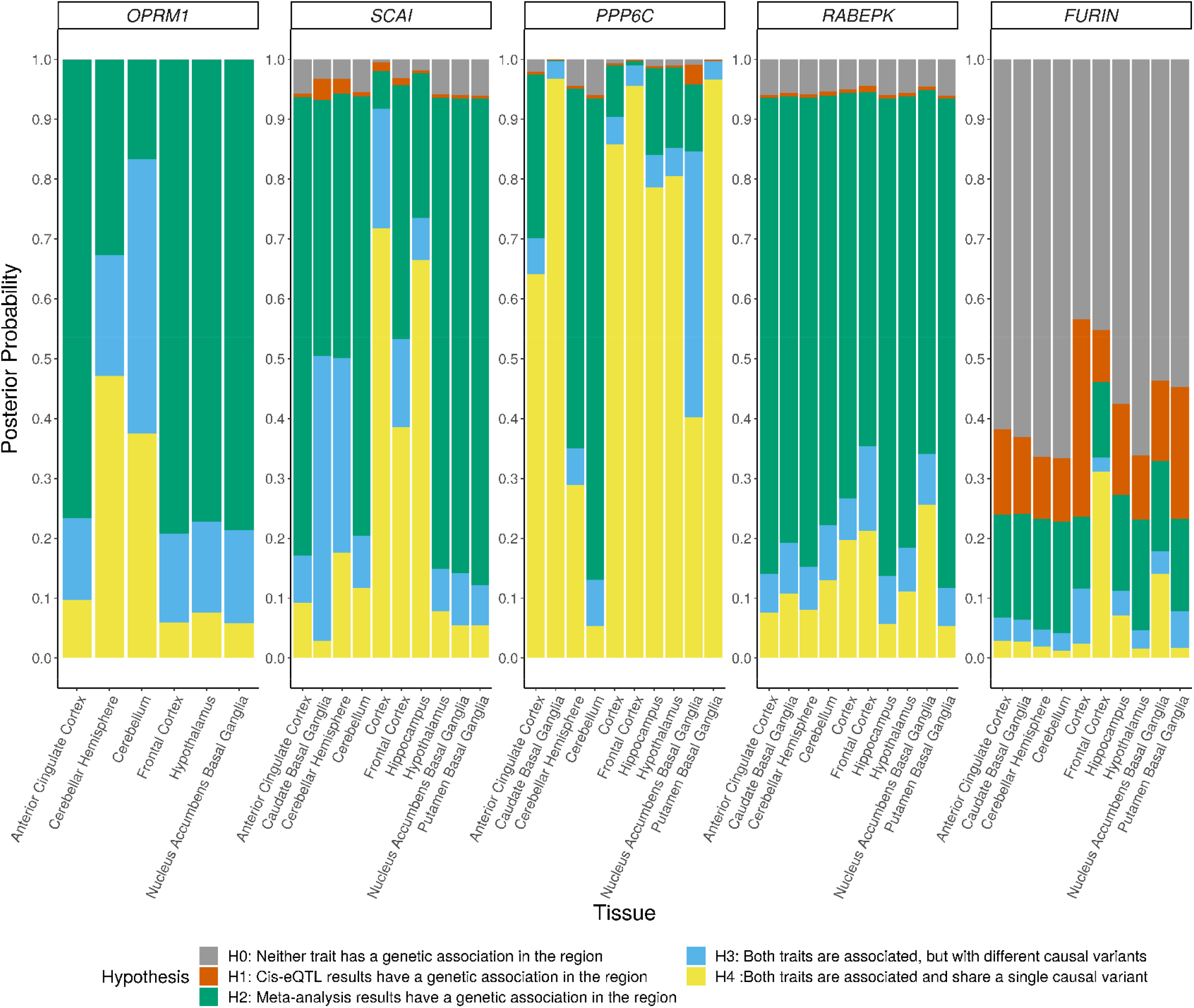
Colocalization of GWAS loci and QTLs for selected genes across 10 brain tissues. Posterior probabilities of supporting hypotheses regarding the association of each trait with SNPs in a region were calculated using coloc. For *OPRM1*, SNP-gene cis-eQTL associations were reported in GTEx Analysis v8 for only 6 of the 10 tissues.

### Drug repurposing analyses suggest druggabillity of all three genes: *OPRM1, PPP6C*, and *FURIN*

To characterize the potential for new pharmacological treatments of OA through drug repurposing or compound development, we examined *OPRM1, PPP6C*, and *FURIN* across multiple drug repurposing databases (the Drug Gene Interaction Database v.3.0 [DGIdb],^42^ Connectivity Map [CMap],^43^ PHAROS [https://pharos.nih.gov/]^44^) *OPRM1* is a known target of more than one-hundred drugs and compounds, including illicit drugs, abused therapeutics (e.g., heroin and oxycodone), and OA treatments (e.g., methadone and buprenorphine)(Supplementary Table 15a-c). In contrast, *PPP6C* is not a target of any known drug or compound but has a 94% likelihood that its protein has ligand properties based on its chemistry.^45^ *FURIN* is the target of one approved drug, pirfenidone, which is indicated for treatment of idiopathic pulmonary fibrosis. There are more than 80 compounds identified targeting *FURIN*, most developed as inhibitors targeting *FURIN* function in infectious diseases: Supplementary Table 16a-c.

### Testing previously reported GWAS variant associations supports three variants: rs1799971, rs62103177, and rs640561

Among the previously reported associations, the strongest association in the current results was observed for rs1799971 (*p*=1.94×10^−6^). The only other previously reported variant tested in our gSEM was rs62103177 in the *KCNG2* gene, which was nominally associated with OA in our EA cohort (*p*=0.0024), though its initial report was among AA only and we were unable to test for replication in AAs owing to a lack of available results that are independent of the initial study.^46^ The GENOA AA analyses are not independent of the Yale-Penn cohorts in the original study. We extended our lookup of previously reported variants to the standard logistic regression meta-analysis we ran with the subset of EA GWAS cohorts without overlapping participants for previously reported EA specific findings. The *CNIH3* variant, rs10799590, and the PGC-SUD variant, rs201123820, were not statistically significant in our standard meta-analysis (p=0.49 and p=0.63, respectively). Of the two PH reported variants, only rs10014685 was present in independent cohorts, but it was not significant in either (deCODE p=0.89; UHS p=0.36). Examining a recently reported GWAS of prescription opioid misuse (POU),^47^ we see a moderate genetic correlation between OA and POU (r_g_=0.74, *p*=2.24×10^−12^) and extend their association of rs640561 to OA (rs640561-T, beta = -0.061, *p*=0.009). Finally, we examined our gene-based GWAS results for evidence supporting previously reported genes and found no support for *GRM8* (*p*=0.591) or *CNIH3* (*p*=0.184), but nominal support for *BEND4* (*p*=0.0023) association with OA, which was reported as genome-wide significant in the PGC-SUD GWAS for the opioid use phenotype (exposed vs. unexposed controls)^26^ and *PTPRF* for POU (*p*=0.026).^47^

## Discussion

Opioid misuse, addiction, and overdoses remain at crisis levels in the United States. Identification of new genetic drivers of OA phenotypes could lead to much needed new pharmacological treatments. In this study, we demonstrated a high degree of genetic correlation between differing diagnostic and frequency-based case definitions of OA and across different types of controls (opioid exposed, unexposed, and population controls), which allowed us to conduct the GENOA meta-analysis and apply gSEM successfully to GENOA and existing summary statistics to conduct the largest GWAS among European ancestry cohort participants to date (23,367 cases and total effective sample size of 88,114 individuals) with the OA case definition. The GENOA GWAS identified European ancestry specific genome-wide significant associations (rs28386916) but the variant was not available in the MVP or PH cohorts and, consequently, was not tested for replication nor was the variant present in the gSEM GWAS. In the gSEM GWAS we found the strongest statistical evidence to date linking variants in intron 1 of the *OPRM1* gene to OA, extending previous candidate gene studies focused on this gene.^37,48,49^ Haplotype analysis of this locus indicated that the long studied and GWAS-identified variant rs1799971 (*OPRM1*-A118G)^27^ may not be the driving variant for this locus’s association with OA. Gene-based analyses of these combined data also identified two novel genome-wide significantly associated genes for OA: *PPP6C* and *FURIN*. Examining the predicted differential expression of these genes in brain tissue and their colocalization with OA association signals suggest that the effect of the *PPP6C* locus on risk of OA is likely to be through effects on *PPP6C* expression, while the signal for *OPRM1* is more complex; there is limited evidence that expression differences explain the association of *FURIN* with OA.

The eight previously published GWAS of OA phenotypes have yielded inconsistent results. The first two used small samples and numbers of variants: N=205 European Americans (EAs)^21^ and N=775 EAs and AAs.^22^ No SNPs met genome-wide significance, but different variants in a glutamate receptor gene (*GRM8*) were among the top candidate gene variants in both studies.^22^ The next two larger GWAS found genome-wide significant associations in two different genes: (1) a potassium channel gene (*KCNG2*) associated with number of opioid dependence symptoms among users (total N=5,432 AAs and *p*=3.6×10^−10^ for rs62103177),^46^ and (2) SNPs in a regulator gene of the glutamate system (*CNIH3*) associated with opioid dependence (total N=2,637 of European ancestry and smallest *p*=4.3×10^−9^ for rs10799590).^24^ The most recent and largest OA GWAS to date are from the PGC-SUD, ^26^ MVP, ^27^ and PH,^28^ which were incorporated in our gSEM GWAS. The PGC-SUD examination of opioid use disorder (OUD; N=4,503 OUD cases, 4,173 opioid-exposed controls) found no genome-wide significant associations; the analysis of OUD vs unexposed controls among AAs (N=1,231 cases, 6,111 controls) found one genome-wide significant association (rs201123820 [*p* = 2.9 × 10^−8^]) but no clear connection to addiction. The MVP tested for variant associations with OUD cases and opioid-exposed controls, finding one genome-wide significant association (*p*=1.51×10^−8^) for rs1799971 in *OPRM1*. Genome-wide significance was achieved by combining the MVP EA cohort with Yale-Penn and SAGE EA cohorts for a combined sample of 10,544 cases and 72,163 controls. PH tested variant associations among EAs with OUD cases defined by ICD9 and ICD10 codes (N=1,039) compared to both general controls without substance use disorder diagnoses (N=20,271) and a subset that also had a medical record of having been prescribed an opioid (N=10,744). Analyses using both control groups identified a genome-wide significant locus on chromosome 4 (rs10014685 [*p*=2.40×10^−8^ using all controls and *p*=1.75×10^−9^ using exposed controls]), while the exposed control analysis found an additional locus on chromosome 16 (rs12931235 [*p*=7.18×10^−10^]). OUD associations for these loci, both intergenic and of unclear functional relevance to OUD. Testing these previously reported variants in the gSEM and standard meta-analysis GWAS results found support for three variants, rs1799971, rs62103177, rs640561. Although the original association between rs62103177 and opioid dependence was found among AAs only, our gSEM results extend that association to EAs. We were unable to test for replication in AAs due to a lack of independent AA GWAS results. rs62103177 is an intronic variant in the potassium voltage-gated channel modifier subfamily G member 2 (*KCNG2*) gene, which has been suggested to have a role in substance use disorders.^23,50^ In GTEx, this variant is also an expression quantitative trait locus in brain tissues for the nearby RBFA Downstream Neighbor (*RBFADN*) gene (*p*<6.1×10^−6^ across 12 GTEx brain tissues). Additional evaluation of this variant and its function in brain are supported by these results.

The top finding of this gSEM GWAS for OA was centered in intron 1 of the *OPRM1* gene (lead SNP rs9478500-C, beta = 0.136, *p*=2.56×10^−9^). Prior candidate gene studies of this region have found nominal associations of variants in intron 1, including some of those that are genome-wide significant here (e.g., rs1381376, rs3778151, & rs3778150).^37,48,49^ As the mu-opioid receptor gene, *OPRM1* has long been a target of OA research and drug development. The functional coding variant rs1799971 (A118G), encoding the amino acid change Asn40Asp, has been studied at length with equivocal results.^37,51,52^ In the current GWAS era, only the MVP GWAS of OUD found rs1799971 to be genome-wide significant (*p*=1.51×10^−8^).^27^ Adding cohorts to the MVP summary statistics in the current study reduced the variant’s association with OA to *p*=1.94×10^−6^, which may be due to variation in haplotype prevalence between studies (discussed below), increase in phenotypic heterogeneity (despite high overall genetic correlations among OA phenotypes), or stochastic variation.

Following prior candidate gene studies,^37,38^ we examined the associations of specific *OPRM1* haplotypes with OA. Three haplotypes were formed by rs1799971 and the genome-wide significant variants: (A) a haplotype with major alleles at all variants; (B) a haplotype with the minor rs1799971-G allele and major alleles at all other variants; and (C) a haplotype with the major rs1799971-A allele and minor alleles at all other variants. In this analysis, the OA association was strongest with haplotype C (*p*=2.43×10^−9^), which was associated with increased risk compared to both haplotype A and haplotype B. Haplotype B, which carried the rs1799971-G allele, was strongly not significant (p=0.52). Evaluation of haplotype associations with OA in this study were limited to a subset of the cohorts for which we had raw genotype data. In this subset, the variant level association for rs1799971 was also not significant (p=0.135), which limits the strength of our conclusions. However, our earlier haplotype analyses of *OPRM1* intron 1 variants and rs1799971 came to similar conclusions, albeit in more limited datasets.^37,38^ This relationship between the underlying EA haplotype structure and risk for OA may explain the equivocal findings at the individual rs1799971 variant level, but it may be that other unidentified variants could be the true causal variants driving these haplotype associations.

The role of genetically driven *OPRM1* expression also appeared complex in this study. In our S-PrediXcan analysis, we observed statistically significant, predicted differential expression of *OPRM1* for OA in cerebellum, the brain tissue with the highest level of *OPRM1* expression in GTEx. Moreover, one of the two *cis*-eQTL variants in the version 8 GTEx model for *OPRM1* expression (rs478498) is in high LD with our top association variant (rs9478500; r^2^=0.56, D’= 0.98), suggesting that the intron 1 locus may have its effect on OA through *OPRM1* expression. However, the colocalization analysis was more equivocal. The hypotheses with the greatest posterior probabilities were that both *OPRM1* expression and OA risk are associated with this locus, but with different causal variants (H3, *posterior probability*=0.46) and with a single causal variant (H4, *posterior probability*=0.38). Given the generally low level of *OPRM1* expression across bulk brain tissues, larger sample sizes and single nuclei experiments will be needed to further distinguish which of these hypotheses is most likely. Ultimately, model organism or organoid experiments are likely to be necessary to fully test gene expression as a potential mechanism for the association of this locus with OA.

Beyond *OPRM1*, we also observed a genome-wide significant association with OA for the intergenic variant rs13333582. Variants in high LD had much weaker associations (*p*=2.25×10^−3^), which may indicate that the rs13333582 association with OA was a false positive. However, rs13333582 is an eQTL for *RANBP10* in multiple brain tissues (Supplementary Table 17). *RANBP10* has been associated with regulation of dopamine D_1_ and mu-opioid receptors^53^ and was recently linked with the variant rs8052287 and substance use disorder— gene interactions that included opioids.^54^ LD between rs13333582 and rs8052287 is moderate in European ancestry (r^2^=0.60, D’=0.81). However, the association between rs8052287 and OA in this gSEM GWAS was only nominally significant (beta=-0.083, *p*=0.0239).

Increasing statistical power through a gene-based GWAS of the gSEM summary statistics identified two new genome-wide significant genes for OA: *PPP6C* and *FURIN. PPP6C* (Protein Phosphatase 6 Catalytic subunit gene) is a component of a signaling pathway regulating cell cycle progression known to be involved with the immune system and cancer(https://www.uniprot.org/uniprot/O00743#function) However, the gene is also strongly expressed across adult human brain tissues (Supplementary Figure 17c) and is linked to abnormal locomotor behavior in mice (http://www.informatics.jax.org/diseasePortal/genoCluster/view/20628).^55^ Predicted biological processes for *PPP6C* include G-protein coupled purinergic nucleotide receptor signaling pathway (GO:0035589) (https://maayanlab.cloud/archs4/gene/PPP6C), which affects regulation of neurons, microglia and astrocytes ^56^. Predicted genetically driven differential expression of *PPP6C* by OA was significant across a number of brain regions, and colocalization analysis of *PPP6C cis*-eQTLs and the OA-variant association signal at this locus also showed high probability of being driven by a shared single variant. Because the *PPP6C*-centered association locus extends into the nearby genes *SCAI* and *RABEPK*, and significant predicted genetically driven differential expression of *SCAI* was also observed, we cannot exclude the possibility that these other genes play a role in, or are responsible for, the *PPP6C*-OA association. However, the degree of gene expression/variant association colocalization for *PPP6C* across brain tissues suggest it as the leading candidate for follow-up studies.

The genome-wide significant gene-based association of OA with *FURIN* was driven by a single variant, rs17514846. However, this signal was supported by analysis of additional *FURIN* variants in a subset of cohorts where more *FURIN* variants were available, including a genome-wide significant association with OA at the variant level for rs11372849. *FURIN* (Furin, Paired Basic Amino Acid Cleaving Enzyme gene) is a member of the convertase family and encodes a type 1 membrane bound protease that is expressed in neuroendocrine and brain tissues, among others (https://www.ncbi.nlm.nih.gov/gene/5045). Although *FURIN* shows higher expression across brain tissues than *OPRM1*, the S-PrediXcan analysis did not show significant predicted genetically driven expression differences associated with OA for this gene, and the colocalization analysis highest posterior probabilities favored no association of either eQTLs or the OA-variant association signal at this locus. However, this may reflect the single variant association with OA in the gSEM GWAS and an effect on OA through mechanisms other than gene expression.

LDSC analyses demonstrated moderate to strong genetic correlations between OA and a variety of substance use, psychiatric, and cognitive phenotypes in expected directions (e.g., positive correlation with cannabis use disorder, inverse correlation with age of smoking initiation). Focusing from the general genomic signal to the specific OA-associated genes, we observed important differences in *OPRM1, PPP6C*, and *FURIN* associations with brain- and SUD-related phenotypes. Although *OPRM1* has been broadly studied, from a GWAS perspective variants in this gene are specifically associated with OUD in the MVP GWAS^57^ and in a GWAS of methadone dose.^58^ No GWAS of other brain- or SUD-related phenotypes have reported an association with *OPRM1*, as indexed by Open Targets (Supplementary Figure 18; Supplementary Table 14a).^59,60^ The variant associated with methadone dose, rs73568641, was not associated with OA in this gSEM GWAS (*p*=0.328). In contrast with *OPRM1*, variants in *PPP6C* have been associated with numerous brain- and SUD-related phenotypes, notably opioid medication use, alcohol consumption, numerous smoking phenotypes, and depression among others (Supplementary Figure 19; Supplementary Table 14b). Indeed, the specific *PPP6C* variants associated with OA in this gSEM GWAS, albeit at p-values in the 10^−7^ range, have been associated with neuroticism, depressive symptoms and a number of smoking phenotypes at genome-wide significance (Supplementary Table 14b). Variants in *FURIN* have been associated with other brain- and SUD-related phenotypes, most predominantly schizophrenia, but also risk taking, number of sexual partners, and insomnia (Supplementary Figure 20; Supplementary Table 14c). The variant driving the genome-wide significant gene-based result here (rs17514846) and the genome-wide significant variant in our subset meta-analysis for OA (rs11372849) are associated with each of these Open Target-identified phenotypes at genome-wide significance (Supplementary Table 14c).

Similar variability was seen across *OPRM1, PPP6C*, and *FURIN* in drug repurposing analyses. While *OPRM1* is the known target of more than one-hundred drugs and compounds, *FURIN* is the target of one approved drug (pirfenidone), and PPP6C is not a target of any known drug or compound across the databases evaluated: DGIdb^42^, CMap^43^ and PHAROS.^44^ However, *FURIN* is the target of more than 80 compounds and the PPP6C protein has a 94% likelihood of being ligandable. Should these novel gene associations be validated in subsequent studies they appear to be potentially important drug development targets.

Among this study’s limitations, the most notable is the focus on cohorts of European ancestry. This focus was required to maximize sample size and statistical power by combining summary statistics across GENOA, PGC-SUD, MVP and PH GWAS through application of the gSEM GWAS method. This approach allowed us to more than double the number of cases used in the EA-focused MVP-SAGE-YP analyses that yielded genome-wide significance for rs1799971 (n=23,367 vs n=10,544) by leveraging gSEM’s ability to model multiple correlated phenotypes and account for sample overlap. These gSEM analyses will be extended to African Americans when the needed ancestry-specific LDSC reference panel becomes available. An additional potential limitation is the variability in OA case definitions (e.g., diagnostic and frequency of use) and types of controls (e.g., exposed, unexposed, and population-based) used to define OA phenotypes across the cohorts within the GENOA and across the other contributing GWAS. However, the genetic correlations across phenotypes were uniformly high (*r*_*g*_>0.9) and resulted in a well-fitting single latent factor gSEM model. An important caveat to the high correlations observed across cohorts with exposed, unexposed, and population controls is that the exposure to opioids was often based on prescribed medication (MVP and PH), which differs in risk of OA from exposure to illicit heroin use. Fine-grained comparison of large samples with different types of exposure to opioids will be needed to resolve this question. Because we have incorporated GENOA and previously published GWAS of OA for our discovery analyses, we do not have independent replication cohorts available in which to test the identified associations. However, OA associations with the intron 1 locus have been previously reported,^29,37,48,49^ the chromosome 16 (rs13333582) is an eQTL for a gene previously reported as associated with OA,^54^ and variants in both *PPP6C* and *FURIN* have previously been associated with substance use and psychiatric disorder traits that are highly associated with OA (Supplementary Figures 12 and 13). Thus, independent replication remains to be demonstrated, but the available evidence supports the identified variants and genes as associated with OA.

In this study, we leveraged gSEM across new and existing OA GWAS that employed various OA phenotypes to conduct the largest EA-focused GWAS to date. Our results show the strongest statistical evidence to date for an association between variants in intron 1 of *OPRM1* with OA. Haplotype analysis of the genome-wide significant variants and previously associated rs1799971 (A118G) suggest that it is the intron 1 variants rather than rs1799971 that is responsible for this association signal, although other unidentified variants outside the tested haplotypes could explain the observed results. Gene-based analyses identified two genome-wide significant associations: *PPP6C* and *FURIN*. These genes are novel for OA, however, variants within them have been associated at genome-wide significance with related phenotypes, such as cigarette smoking, alcohol consumption, general risk taking, and schizophrenia. With strong SNP-based heritability for these OA phenotypes, but only these few genome-wide significant findings, it is clear that increased sample sizes for GWAS and complementary approaches (e.g., gene regulation in postmortem brain tissues) are needed to identify much of the genetics driving risk of OA as well as to extend these studies to non-European ancestries.

## Methods

### Cohorts and Opioid Addiction Phenotype

Descriptive statistics for the GENOA studies contributing previously unpublished GWAS of opioid addiction (OA) to this investigation are provided in Supplementary Table 1, and full descriptions of the studies are provided in the Supplementary Methods. In total, this analysis provides new OA GWAS results for 304,831 individuals, including 7,281 cases and 297,550 controls, with an effective sample size of 88,114 (4 / ((1 / # Cases) + (1 / # Controls))).^61,62^

In the GENOA studies, OA was defined either based on frequency of opioid use (FOU) or Diagnostic and Statistical Manual (DSM) of Mental Disorders criteria. Some studies included only opioid-exposed individuals in their control groups, while others included both exposed and unexposed individuals.

### Genotype Quality Control and Imputation

Sites in the GENOA consortium conducted standard genotype quality control using filters appropriate for their samples. SNPs were filtered based on call rate and deviation from Hardy-Weinberg equilibrium. Samples were filtered based on call rate, excessive homozygosity, relatedness, and sex discrepancies. Classification of European ancestry individuals was based on comparison to reference populations using STRUCTURE ^63^. Specific filters used for each sample are provided in Supplementary Table 18.

For most samples, genotype imputation was performed with the Michigan Imputation Server ^64^ using the 1000 Genomes Phase 3 v5 reference panel. For COGA, genotypes were phased with SHAPEIT2^65^ and imputed with Minimac3^64^ using the 1000 Genomes Phase 3 v5 reference panel. For deCODE, genotype imputation was conducted by long-range phasing and haplotype imputation of chip-genotyped individuals with methods described previously ^66^.

### Association Testing

Imputed genotypes for GENOA studies were tested for association with opioid addiction case-control status using rvtests ^67^, adjusting for sex, age, genotype principal components and in some cases recruitment site and other study-specific covariates. deCODE data were analyzed using logistic regression treating disease status as the response and imputed genotype counts as covariates. Other available individual characteristics that correlate with disease status were also included in the model as nuisance variables (sex, age, county of origin(ref1), blood sample availability, and an indicator function for the overlap of the lifetime of the individual with the time span of phenotype collection) using previously described methods.^68^ To account for inflation due to population stratification and relatedness, test statistics were divided by an inflation factor (1.10) estimated from linkage disequilibrium score regression (LDSR).^69^ Inverse variance-weighted meta-analysis of chromosome 15 variants with MAF<0.01 and Rsq<0.8 in the region containing *FURIN* was performed using METAL ^70^ including only studies with no overlapping samples (GENOA + MVP without SAGE and Yale-Penn + Partners; Cases N = 16,849, Controls N = 379,493, Total N = 396,342, Effective Total N = 52,508).

### Cohort Descriptions for GenomicSEM

The gSEM analysis includes results on EA from the GENOA meta-analysis, Million Veteran Program (MVP), Psychiatric Genetics Consortium Substance Use Disorders Group (PGC-SUD), and the Partners Health cohorts. The MVP results are based on a meta-analysis including MVP parts 1 and 2 (total of 8,529 OUD cases and 71,200 opioid-exposed controls), along with Yale-Penn and Study of Addition: Genetics and Environment (SAGE) cohorts (total of 10,544 OUD cases and 72,163 opioid-exposed controls). ^27^ The PGC-SUD results include 4,503 opioid dependence cases and 4,173 unexposed controls. ^26^ Unexposed controls are used for the PGC-SUD because the exposed controls results have negative heritability estimates. The partners health cohort includes 1,039 OUD cases and 10,743 exposed controls. ^28^ Note that the GENOA GWAS, Yale-Penn, and SAGE parts of the MVP, and the PGC-SUD results include overlapping samples. However, accounting for this sample overlap is a feature of the gSEM approach applied in this study. A total of 2,434,903 variants were present in all cohorts and tested for association with the OA latent variable in the gSEM analysis using a total sample size of 403,915 (23,367 cases and 384,619 controls; effective sample size of 88,115).

### Genomic Structural Equation Modeling

The GenomicSEM package ^30^ within R was used for genomic structural equation modeling (gSEM). The gSEM implemented multivariable LDSC function within this package was used to calculate single-nucleotide polymorphism heritability on the observed and liability scale (prevalence of 10%), genetic covariance matrices, and genetic correlation. The LD scores from 1000 Genomes Project phase 3 European ^69^ were used as the reference population in this calculation. The sampling genetic covariance matrix is expanded to incorporate SNP effects by including the covariances between SNPs and each cohort. This expanded sampling genetic covariance includes the multivariate LDSC estimated genetic variances and covariances, along with the sampling covariance matrix of the SNP effects on the cohorts, which are estimated using cross-trait LDSC with the sampling correlation weighted by the sample overlap. With the gSEM implemented LDSC, the overlap of samples between GENOA, MVP meta-analysis, and PGC-SUD is not a concern.

A single latent factor gSEM was used with the residual variance of the latent factor set to 1 to normalize the loading estimates. The loadings were calculated using diagonally weighted least squares and residual variances were bound to above 0.01 to avoid negative residual variance estimates.

### Haplotype Analyses

To conduct haplotype analyses raw genotype data is needed. Chromosome 6 genotypes were available from UHS, VIDUS, ODB, Yale-Penn, CATS and Kreek (Supplementary Table 1). These cohorts’ data were phased with Eagle v2.4 via the Michigan Imputation Server ^64^. Haplotypes for samples from each study were constructed by extracting *OPRM1* variants that were genome-wide significant in the gSEM analysis and concatenating ordered by genomic position. Supplementary Table 19 provides counts for the various haplotypes observed across the studies. The 3 most common haplotypes, which accounted for 98% of observed haplotypes, were tested for association with OA in R adjusting for sex and genotype principal components. Only individuals carrying exclusively these haplotypes were included in the analysis. Two models were run, one in which the haplotype containing all major alleles served as the reference haplotype and one in which the haplotype containing the minor rs1799971-G allele served as the reference. This approach provided three effective comparisons: (a) rs1799971-G haplotype versus the major allele haplotype; (b) minor allele + rs1799971-A haplotype versus the major allele haplotype; (c) minor allele + rs1799971-A haplotype versus rs1799971-G haplotype. Individual cohort results were combined in an inverse variance-weighted meta-analysis using METAL (N = 21,037).

### Gene-Based Analyses

Gene-based associations with OA were calculated from the gSEM summary statistics with MAGMA v1.08 ^39^ with a 10 kb gene window via the Functional Mapping and Annotation (FUMA) of GWAS web tool v1.3.6a ^71^. The gSEM summary statistics were mapped to 15,977 protein coding genes, resulting in a Bonferroni-corrected threshold of p = 3.129e-6 for declaring genome-wide significance.

### Cross-trait genetic correlations with OA

Summary statistics from the gSEM were used as input into LD score regression (LDSC) with reference to the 1000 Genomes EUR panel to estimate genetic correlations between OA and 38 other complex phenotypes. These phenotypes were categorized into the following groups: drug and alcohol use, cigarette smoking, psychiatric, personality, neurological, cognitive/education, and brain volume. The full list of phenotypes and GWAS datasets, as obtained from LD Hub or shared by the original investigators, are provided in the Supplementary Table 20.

### PrediXcan

To investigate the transcriptome-wide associations between predicted gene expression and OA, we employed the MetaXcan v0.6.6 ^40^ method. Briefly, MetaXcan uses association summary statistics to predict associations between gene expression and a phenotype of interest association. Gene expression models were predicted from tissue-specific eQTL datasets. To increase the performance of our prediction models, we used the MASHR-M ^72^ models built on fine-mapped variables from DAP-G.^73^ The specific models we used were pre-computed MetaXcan models available through PredictDB (http://predictdb.org/) for 12 brain regions (Supplementary Table 21) that were generated using the GTEx ^74^ version 8 datasets.

Summary level statistics from the Genomic SEM analysis were used as input to MetaXcan. Prior to input, summary statistics were harmonized according to the best practices guide outlined on the MetaXcan wiki. As part of this process, the gwas_parsing.py utility (https://github.com/hakyimlab/summary-gwas-imputation) was used to lift summary statistics over to the human genome build version 38 and provide harmonized variant identifiers compatible with those used by GTEx v8. To increase the number of overlapping markers between our summary statistics and the fine-mapped pre-built MASHR models, we imputed missing summary associations as suggested by the best practices workflow. Imputation was performed separately for each chromosome using the gwas_summary_imputation.py utility (https://github.com/hakyimlab/summary-gwas-imputation) and the pre-computed parquet genotype, genotype metadata files, and European LD block files available through the MetaXcan zenodo repository ^75^. Imputed summary statistics were finally re-combined using the gwas_summary_imputation_postprocess.py utility.

The resulting imputed, harmonized association summary statistics were then used as input to MetaXcan. The number of genes tested for each tissue is found in Supplementary Table 21. FDR correction was applied to account for the number of genes tested across all tissues (156215 total tests). A gene’s predicted expression was considered significantly associated with OA if its FDR-adjusted p-value fell below a threshold of 0.05.

### Colocalization

Co-localization analysis was performed using the coloc package in R.^76^ Cis-eQTL data for individuals of European ancestry from the GTEx v8 eQTL Tissue-Specific All SNP Gene Associations dataset (dbGaP Accession phs000424.v8.p2) were input as a quantitative trait into coloc (sample sizes for each tissue type indicated in Supplementary Table 22). Summary statistics from the gSEM analysis were input as a quantitative trait, with a sample size of 403,915. Summary statistics from the standard meta-analysis of OA were input as a case-control trait with 16,849 cases and 379,493 controls. All SNP positions were lifted over to build 38. The cis-eQTL data was partitioned into blocks based on the gene in the SNP-gene pair. For each gene block, only SNPs in the gSEM or meta-analysis summary statistics overlapping with the cis-eQTL data were input into the coloc function for (approximate) Bayes Factor colocalization analysis.

## Data Availability

The GWAS summary statistics generated and/or analyzed during the current study will be made available via dbGAP; the dbGaP accession assigned to the UHS is phs000454.v1.p1. The website is https://www.ncbi.nlm.nih.gov/projects/gap/cgi-bin/study.cgi?study_id=phs000454.v1.p1

## Acknowledgements

The late Dr. Mary Jeanne Kreek from the Laboratory of the Biology of Addictive Diseases at the Rockefeller University, New York, NY, was a key contributor to the research presented in this manuscript.

We thank deCODE genetics for contributing summary statistics from their analysis of opioid addiction in the deCODE cohort. We particularly thank Kari Stefansson, Thorgeir Thorgeirsson, Valgerdur Runarsdottir, Thorarinn Tyrfingsson, Gudmundur Einarsson, and Daniel F. Gudbjartsson for their work to generate and share these results.

