## Supplemental Figures for "Multi-trait genome-wide association study of opioid addiction: *OPRM1* and Beyond"

### Supplementary Information

### Supplementary Methods

#### Study Descriptions

Addictions: Genotypes, Polymorphisms, and Function/Human Genetic Correlates of Addictive Diseases. Drug addiction continues to be a major medical and social problem. It is estimated that one million or more persons in the United States are currently addicted to heroin or prescription opioids, with millions more worldwide. Cocaine addiction and alcohol dependence are frequent comorbid conditions in persons with heroin/opioid dependence in addition to being major primary addictions. Many studies over the past thirty years have shown that these drugs disrupt physiologic systems, and that these disruptions may contribute to drug addiction and alcohol dependence and to relapse to drug or alcohol abuse following withdrawal and abstinence. Clinical observations suggest that individuals differ in their response to heroin, cocaine, and alcohol; however, little is known about specific underlying hereditary genetic factors which might influence individual susceptibility to the addictive properties of these substances. Studies also suggest that both common and distinct heritable factors account for the genetic variance in the susceptibility to the separate addictive diseases. We hypothesize that there is a heritable as well as environmental basis for the acquisition and persistence of, and relapse to, specific addictive diseases. Using samples from individuals without and with opioid and other specific drug dependence diagnoses and psychiatric comorbidities, genetic analyses will be used to determine association and linkage. All study subjects will be extensively characterized with respect to the addictive diseases, medical history, family medical addictive disease history; psychiatric comorbidity, and psychological profile, as well as ethnic/cultural background. A better understanding of the consequences of genetic contributions with respect to protection from, or susceptibility to, heroin/opioid addiction and related codependencies and comorbid conditions, could have enormous importance in both prevention and treatment of this problem. dbGaP accession number: phs001109.v3.p2

CaTS2. Participants in the two CATS comparisons (i.e., CaTS2-Mole and CATS-Pert-Hunt) were ascertained from the following four datasets. (1) The Comorbidity and Trauma Study (CATS) is a retrospective case-control study examining genetic and environmental factors contributing to heroin dependence liability. The study was a collaboration of investigators at Washington University School of Medicine, the Queensland Institute of Medical Research, and the National Drug and Alcohol Research Centre at the University of New South Wales. Cases were recruited from clinics providing opioid substitution therapy in the greater Sydney region of Australia. DSM-IV opioid dependence diagnoses were obtained via administration of a comprehensive psychiatric interview based on the Semi-Structured Assessment of the Genetics of Alcoholism- Australia version, revised. For the current analyses, these individuals (N=1240) were compared to population controls (N=1922) from: (2) The Twin Study of Mole Development in Adolescence, an ongoing investigation of melanocytic naevi. Unassessed parents of these twins, used as population controls, were very likely to have a prevalence of heroin dependence lower than that in the general population. The second case-control comparison included cases from (3) the Western Australia Study on Heroin Dependence which included heroin addicted individuals from the greater Perth region focused on genetic contributions to heroin dependence and response to naltrexone treatment. A DSM-IV diagnosis of heroin dependence was derived via clinical assessment. Participants provided blood samples during their treatment at the Perth Naltrexone Clinic (now named as the Fresh Start Recovery Programme). These individuals (N=721) were compared to population controls (N=1199) from (4) the Hunter Community Study, a population-based cohort study established to assess factors important in the health, well-being, social functioning, and economic consequences of aging.

DNA obtained from blood samples was genotyped using either the Illumina 660W-Quad or 610- Quad BeadChip arrays. Quality control was conducted as follows. Genotyped SNPs were filtered due to deviation from HWE (p < 10-6), MAF (≤0.01), call rate (≤95%), and GenomeStudio genotype quality score (≤0.7). Additional analyses were performed to detect cryptic relatedness among genotyped participants using a pi-hat cut-off of 0.1. Principal components analysis (PCA) was performed with the SmartPCA program in the Eigensoft 3.0 package to identify outliers of non-European ancestry, based their relative distance from the center of the northern European group. Participants who were more than six standard deviations from the centroid of the first two eigenvectors were removed. Genotypes were phased using SHAPEIT and then SNP imputation was performed on the Michigan Imputation Server using the 1000 Genomes Phase 3 reference panel. Cases and some control subjects are available in dbGaP under “A [Genome-Wide Association Study](https://www.sciencedirect.com/topics/neuroscience/genome-wide-association-study) of Heroin Dependence” (accession number phs000277.v2.p1).

COGA. COGA is a large family-based study that recruited alcohol dependent probands from treatment facilities from seven sites in the United States ^[^PMID: 9603606, PMID:12875050]. Their extended families were invited to participate. Additional individuals and their families were recruited from the same communities using a variety of resources. Institutional review boards at all sites approved the study, and all participants provided informed consent. All participants were administered a version of the Semi-Structured Assessment for the Genetics of Alcoholism interview (SSAGA; those aged <18 years were administered a child version, the C-SSAGA) [PMID: 8189735, PMID: 10615721]. Diagnosis was by DSM-IV criteria. Data cleaning, quality control and harmonization have been previously described [PMID: 31090166]. Only EA (n=7,631) individuals were included in these analyses.

deCODE. All Icelandic data were collected through studies approved by the National Bioethics Committee (NBC; License #VSN-18-113) following review by the Icelandic Data Protection Authority. Participants donated blood or buccal samples after signing a broad informed consent allowing the use of their samples and data in all projects at deCODE genetics approved by the NBC. All personal identifiers of the participants' data were encrypted by a third-party system, approved and monitored by the Icelandic Data Protection Authority. The GWAS analysis used as cases subjects diagnosed with opioid use disorders by clinicians at Vogur Hospital, Reykjavik, and controls were population controls (excluding individuals diagnosed treated for substance use disorders).

##### NIDA Genetic Epidemiology of Opioid Dependence in Bulgaria (GEODB). The study was conducted in Bulgaria as a collaboration between the Molecular Medicine Center (Medical University, Sofia), the Bulgarian Addictions Institute (Sofia), the Initiative for Health Foundation (Sofia) and Washington University School of Medicine (Saint Louis, MO, USA).

This is a case-population control study. Participants (N=2,499 interviews; N=1,995 with GWAS data) in our study are primarily active heroin users (78%) aged 18 and over, with a median 8.9 years of daily heroin use (minimum of 12 months). Virtually all participants (99.1%) qualified for a lifetime DSM-IV diagnosis of dependence, symptom endorsement probabilities ranging from 77% to 99.3%; and 61% endorsing all symptoms. 85% were heroin injectors and 97% reported using (or having used) heroin daily. Anonymized, population-representative control samples (no phenotypic data, N = 1,062 with GWAS data) were drawn from the MMC repository. Unlike other studies, where samples are clinically ascertained (e.g., from ORT programs), our participants are primarily untreated.

PTSD, major depression, suicidality, mania, alcohol, nicotine and substance dependence (separating heroin and opioids) were assessed using the MAGIC (Todd et al., 2003, PMID: 14627881), a broad semi-structured psychiatric assessment tool with good reliability and prospective stability.

The onset of (any) illicit drug use in this sample is not particularly remarkable (age~16), but the age of onset of heroin use is much lower than in the USA: 25% by age 16; 50% by age 19, with no differences between ethnicities or gender. This is consistent with findings from other studies in Bulgaria. In contrast, in 2008, SAMHSA estimated an average age of 23.4 for new heroin initiates in the USA. Further, progression from first use to regular use appears to be more rapid, with the majority of participants reporting onset of dependence problems within one year of initiation.

Another unique aspect of this study population are the comparatively low levels of polysubstance use (outside of nicotine and alcohol). This early-use, fast-transition and monosubstance heroin pattern stands in sharp contrast to what is reported in many studies in the Western World. In our study, 47% of Ethnic Bulgarians and 66% of Roma used only opioids, even years after the onset of drug use. 15.4% of Ethnic Bulgarians and 66.2% of Roma reported heroin as the very first illicit drug used. Lifetime, 12.5% of Ethnic Bulgarians and 52.8% of Roma report never having used another illicit drug other than heroin more than five times. While both Roma and Ethnic Bulgarians certainly experiment with other drugs, abuse/dependence rates for these drugs are low in Ethnic Bulgarians (from 8.4% for inhalants to 28% for stimulants) and in the Roma population (from 1.3% for hallucinogens to 11.2% for stimulants). dbGaP accession number: phs001804.v1.p1

Urban Health Study (UHS). Study participants were drawn from the UHS, a serial, cross-sectional, sero-epidemiological study that recruited people who injected drugs (PWIDs) in the San Francisco Bay Area from 1986 to 2005.{Kral, 2003 #88} Eligibility criteria for study entry included injection of an illicit drug in the past 30 days, ability to provide informed consent, and age 18 or older. The current study included participants who reported their ancestral origin as Caucasian (henceforth referred to as European Americans [EAs]). The genotyping procedures have been described before in the context of the full genotyped cohort of HIV-1 cases and controls.{Johnson, 2015 #221} Briefly, stored serum samples that remained after HIV testing were used to extract DNA for use as a genotyping source. Sample restoration was conducted using the Illumina Formalin-Fixed Paraffin-Embedded (FFPE) kit to maximize genomic DNA quality, and the restored DNA samples were genotyped on the Illumina Omni1-Quad BeadChip. Over 60% of the genotyped UHS participants met the Office of National Drug Control Policy definition of heroin abuse (injecting 10+ times in the past 30 days),{Morral, 2000 #92}{Rhodes W., 2000 #93} which is highly correlated with clinical levels of dependence on the Severity of Dependence Scale{Gossop, 1995 #94}{Strang, 1999 #95} and with DSM-IV{Association, 1994 #96} heroin abuse/dependence in analyses of the National Household Survey on Drug Use and Health data (87% positive predictive value; see Hancock et al.{Hancock, 2015 #6}). These UHS participants, who abused heroin an average of 80.9 times in the past month and were very likely dependent on it, are henceforth referred to as opioid addiction cases. The remaining genotyped UHS participants, who were addicted to cocaine or other substances but not addicted to heroin, were not included in the current study. dbGaP accession number: phs000454.v1.p1

Population controls used for comparison to the UHS cases were drawn from the following three cohorts in dbGaP: (1) Genome-Wide Association Study of Parkinson Disease: Genes and Environment ([accession number phs000196.v3.p1](https://www.ncbi.nlm.nih.gov/projects/gap/cgi-bin/study.cgi?study_id=phs000196.v3.p1)); (2) High Density SNP Association Analysis of Melanoma: Case-Control and Outcomes Investigation ([accession number phs000187.v1.p1](https://www.ncbi.nlm.nih.gov/projects/gap/cgi-bin/study.cgi?study_id=phs000187.v1.p1)); (3) National Institute of Diabetes and Digestive and Kidney Diseases (NIDDK) Chronic Renal Insufficiency Cohort Study (CRIC) ([accession number phs000524.v1.p1](https://www.ncbi.nlm.nih.gov/projects/gap/cgi-bin/study.cgi?study_id=phs000524.v1.p1)).

Vancouver People Who Inject Drugs Study (VPWIDS). VPWIDS participants were recruited from community settings (e.g., low-barrier health and social service organizations, harm reduction venues, open drug markets) in Vancouver’s Downtown Eastside neighborhood, an area with widespread illicit drug use and HIV infection. Eligible participants were aged ≥ 18 years, self-reported white ancestry, used any illicit drug via injection at least once in the 30-day period prior to recruitment, and provide written informed consent. VPWIDS participants were also drawn from individuals participating in two community-recruited prospective cohorts of people who use illicit drugs in Vancouver: the AIDS Care Cohort to evaluate Exposure to Survival Services (ACCESS), a prospective cohort of people living with HIV infection who use illicit drugs; and the Vancouver Injection Drug Users Study (VIDUS), a prospective cohort of people at risk of HIV infection who inject illicit drugs. Eligibility requirements for ACCESS included: Aged ≥ 18 years at the baseline interview, use of any illicit drug (not including cannabis, which was illegal during the study period) via any route of administration at least once in the 30 days prior to the baseline interview, and provide written informed consent. Eligibility requirements for VIDUS included: Aged ≥ 18 years at the baseline interview, injection of any illicit drug at least once in the 30 days prior to the baseline interview and provide written informed consent. At baseline and every six months thereafter, VIDUS/ACCESS participants completed an interviewer-administered questionnaire eliciting information on socio-demographics, HIV risk behaviors (if seronegative), HIV treatment patterns (if seropositive), substance use patterns, social/structural exposures, and other relevant exposures/outcomes. They also underwent an examination by a nurse including tests for HIV and hepatitis C virus serostatus (if seronegative) or plasma HIV-1 RNA viral load and other HIV clinical monitoring (if seropositive.) VIDUS/ACCESS participants self-reporting white ancestry and any injection drug use in the 30-day period prior to their most recent study interview were eligible for inclusion in the VPWIDS. The VIDUS, ACCESS and VPWIDS studies were reviewed and approved by the University of British Columbia/Providence Healthcare research ethics board. All VPWIDS participants provided written informed consent. Following recruitment, VPWIDS participants recruited from the community (i.e., not drawn from the VIDUS/ACCESS cohorts): completed an interviewer-administered questionnaire eliciting data on sociodemographics (e.g., sex/gender, age), substance use patterns, and sexual behaviors; underwent a test of HIV serostatus; and if HIV seropositive, plasma HIV RNA-1 viral load and viral genotyping. All VPWIDS participant provided blood specimens for extraction of human DNA and mRNA extraction. VPWIDS participants DNA was genotyped on the Illumina Infinium OmniExpress-24 BeadChip. Opioid addiction was defined as those with heroin or illicit prescription opioid use >= 2-3 times/week in last six months.

Population controls used for comparison to the VPWIDS cases were drawn from the dbGaP deposited study: National Cancer Institute (NCI) Genome Wide Association Study (GWAS) of Lung Cancer in Never Smokers ([accession number phs000634.v1.p1](https://www.ncbi.nlm.nih.gov/projects/gap/cgi-bin/study.cgi?study_id=phs000634.v1.p1)).

Yale-Penn. The Yale-Penn study was designed to conduct genetic studies for addiction using mostly unrelated individuals but also small nuclear families, all of whom were recruited from the eastern United States.39-41 ND was not considered in the inclusion or exclusion criteria, but lifetime FTND data were collected among smokers.42 Genotyping was conducted on the Illumina HumanOmni1-Quad array. QC mimicked prior analysis,7 except that ancestry assignments were refined using K-means clustering to assign individuals based on the nearest centroid across the first 10 PC eigenvectors with reference to 1000G EUR or AFR population. There were 1,579 EAs and 2,637 AAs in the final analyses, which included adjustment for age, sex, and PC eigenvectors.

### Supplementary Note 1

Funding support for each cohort.

ALIVE: The authors thank the participants and staff of the AIDS Linked to the IntraVenous Experience (ALIVE) cohort study. This work was supported by NIDA grants R01DA039408, R01DA047064, and U01DA036297, NIAID K24-AI118591 and T32 DA007292-27

ADAA: R01AA017444

Addictions: The Dr. Miriam and Sheldon G. Adelson Medical Research Foundation; The Clinical and Translational Science Award UL1RR024143 from the National Center for Advancing Translational Sciences of the NIH (B. Coller).

CaTS2: The Comorbidity and Trauma Study was funded by R01DA17305; The Western Australia Study on Heroin Dependence was funded by the Australia Government’s National Health and Medical Research Council (Grant number 513862); 3) The Twin Study of Mole Development in Adolescence (principal investigator [PI]: Nick Martin) was funded by the Australian Government’s National Health and Medical Research Council (Grant number 389891); and 4) Support for the Hunter Community Study has been previously described ([68](https://www.sciencedirect.com/science/article/pii/S0006322315000463?via%3Dihub#bib68)). Lastly, GWAS genotyping services for “A Genome-Wide Association Study of Heroin Dependence” at CIDR, located at The Johns Hopkins University, were supported by NIH contract number N01 HG65403.

COGA: U01AA008401

COGEND: COGEND was supported by grants from the National Cancer Institute (NCI; grant number P01 CA089392, PI: Laura Bierut) and NIDA (R01 DA036583 and R01 DA025888, PI: Laura Bierut), both of the National Institutes of Health (NIH). Genotype data are available via dbGaP as part of the “Genetic Architecture of Smoking and Smoking Cessation” (accession number phs000404.v1.p1) and “Study of Addiction: Genetics and Environment (SAGE)” (accession number phs000092.v1.p1). Funding support for genotyping, which was performed at CIDR, was provided by 1 X01 HG005274-01 and by the NIH Genes, Environment and Health Initiative [GEI] (U01 HG004422). CIDR is fully funded through a federal contract from the NIH to The Johns Hopkins University, contract number HHSN268200782096C. Assistance with genotype cleaning, as well as with general study coordination, was provided by the GENEVA Coordinating Center (U01 HG004446).

deCODE: [ADD FUNDING SUPPORT]

START: K01DA036751, R01DA044015, and R01DA044015

UHS, VIDUS, and SAGE: R01DA044014, R01DA043980, R01DA038632, R33DA027486

### Supplementary Figures

**Supplemental Figure 1.** Manhattan plot of GENOA African American results.

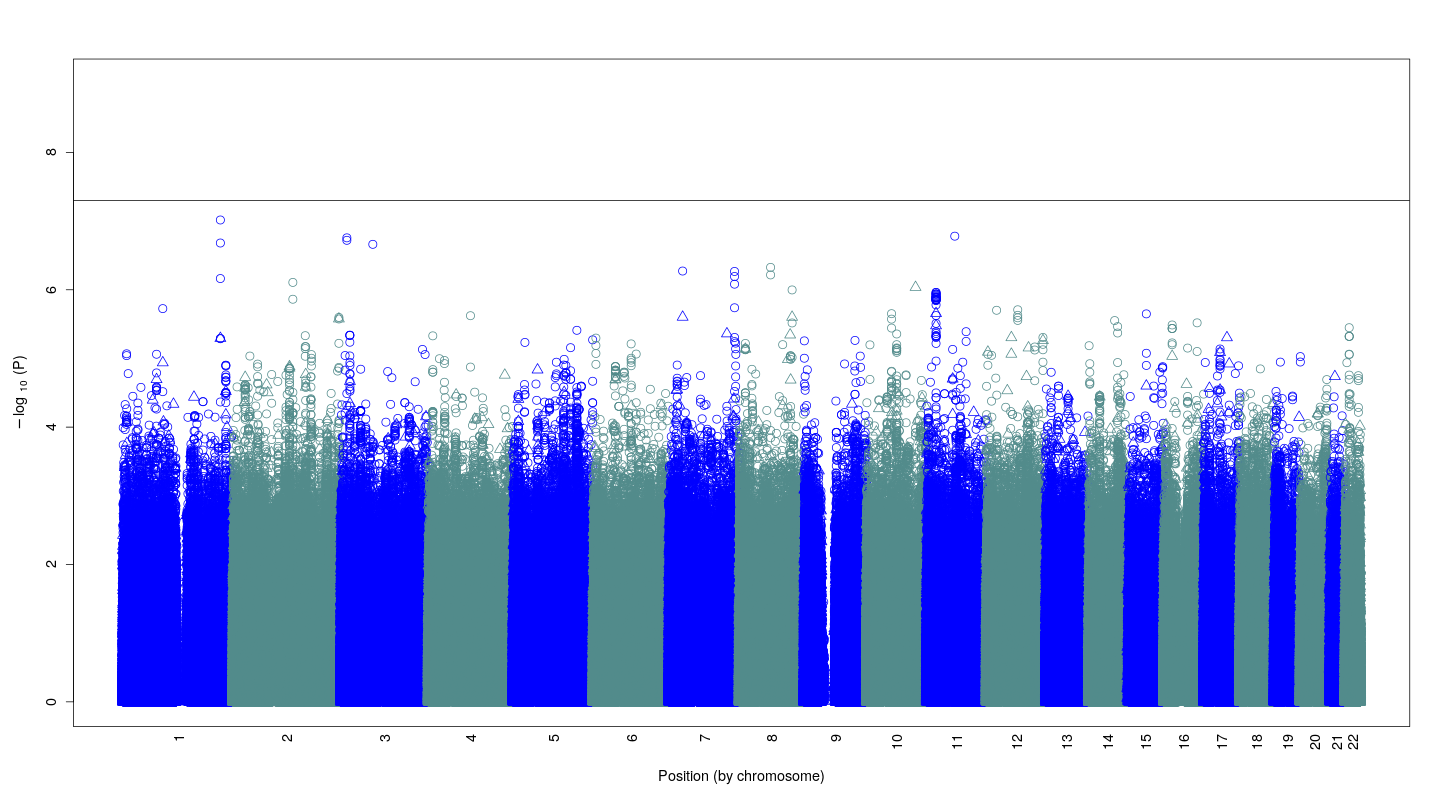

**Supplemental Figure 2.** Quantile-Quantile (QQ-plot) plot of GENOA African American results.

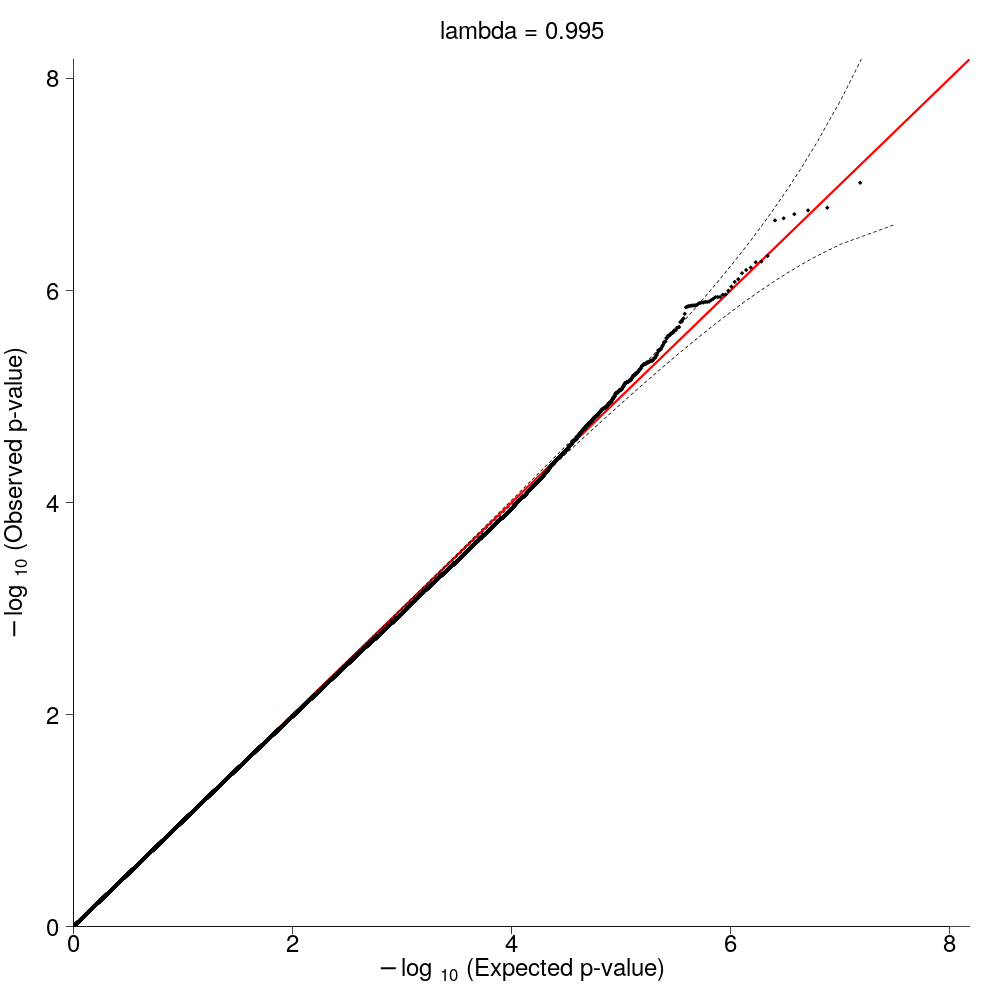

**Supplemental Figure 3.** Manhattan plot of GENOA European American results.

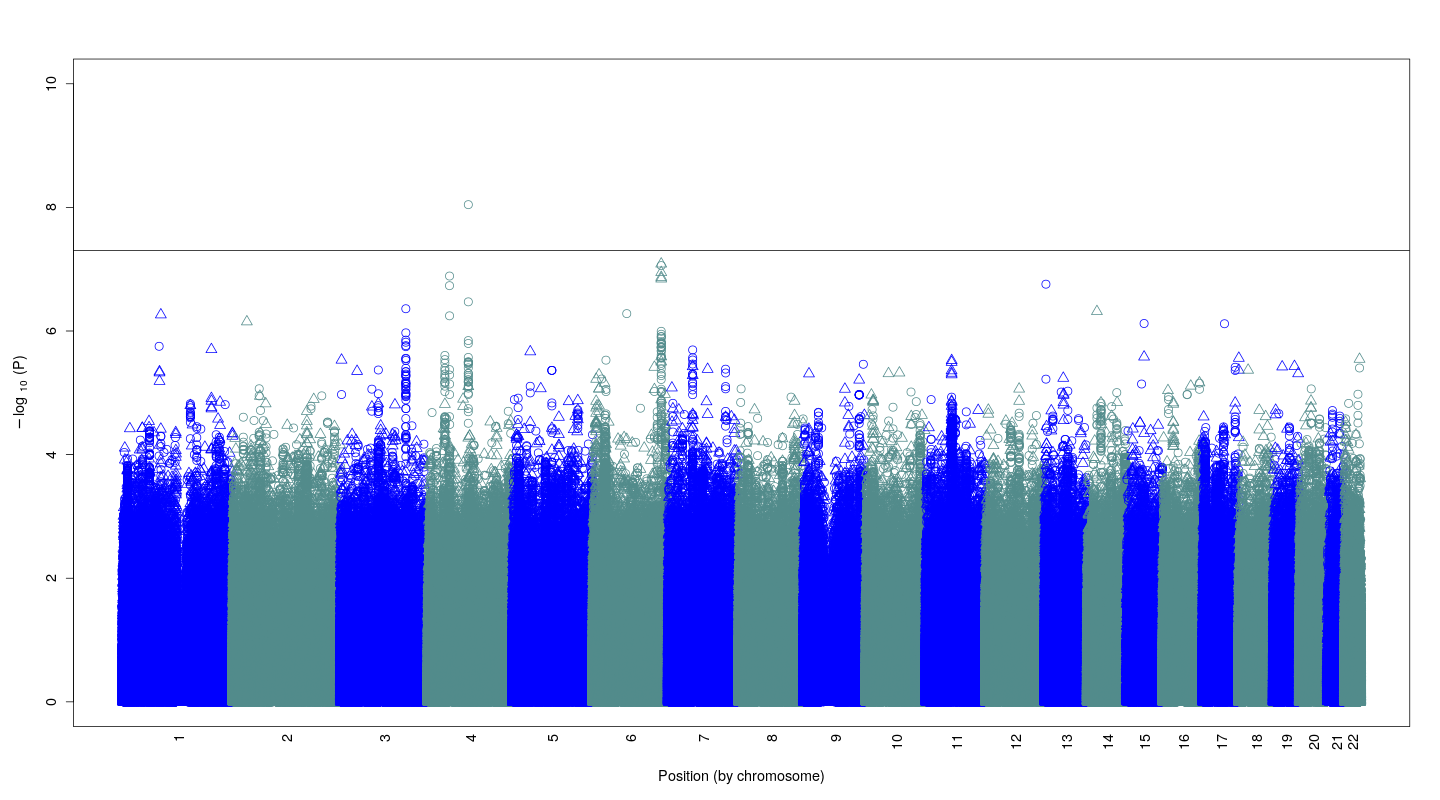

**Supplemental Figure 4.** Quantile-Quantile (QQ-plot) plot of GENOA European American results.

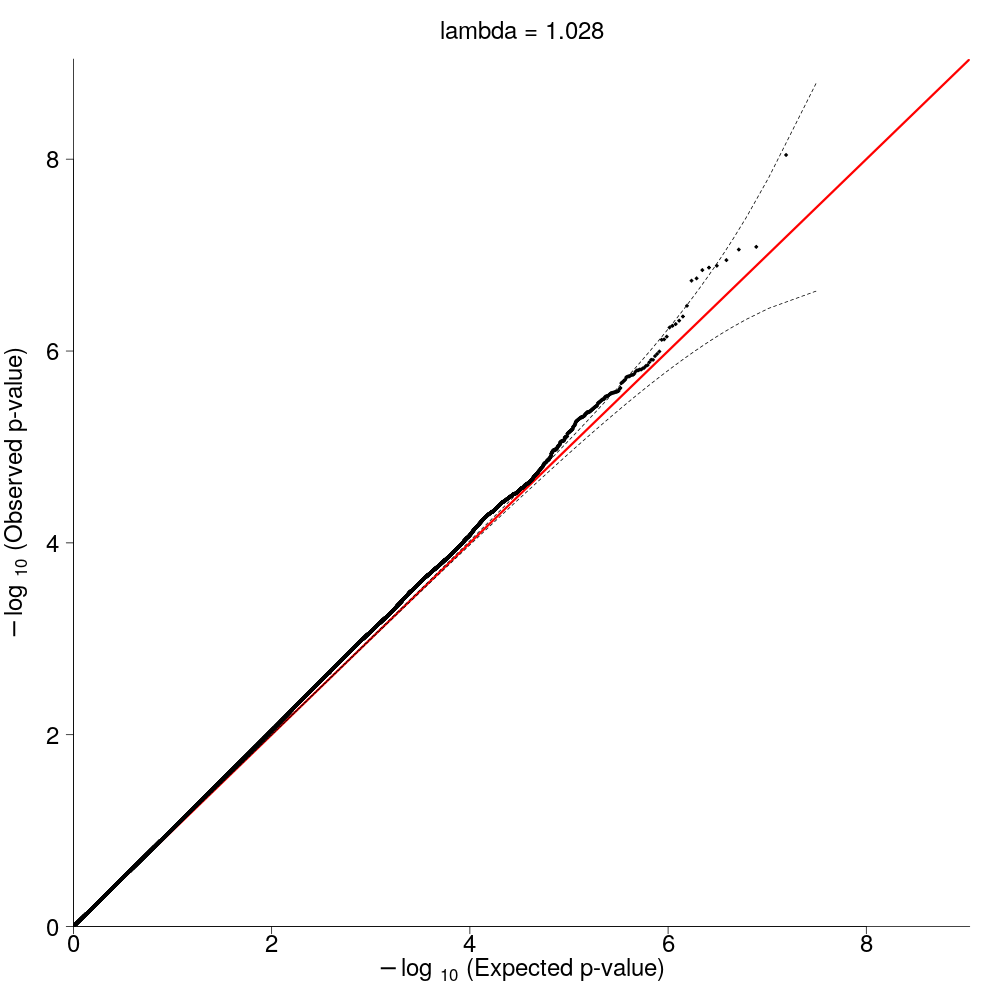

**Supplemental Figure 5.** Manhattan plot of GENOA African American + European American results.

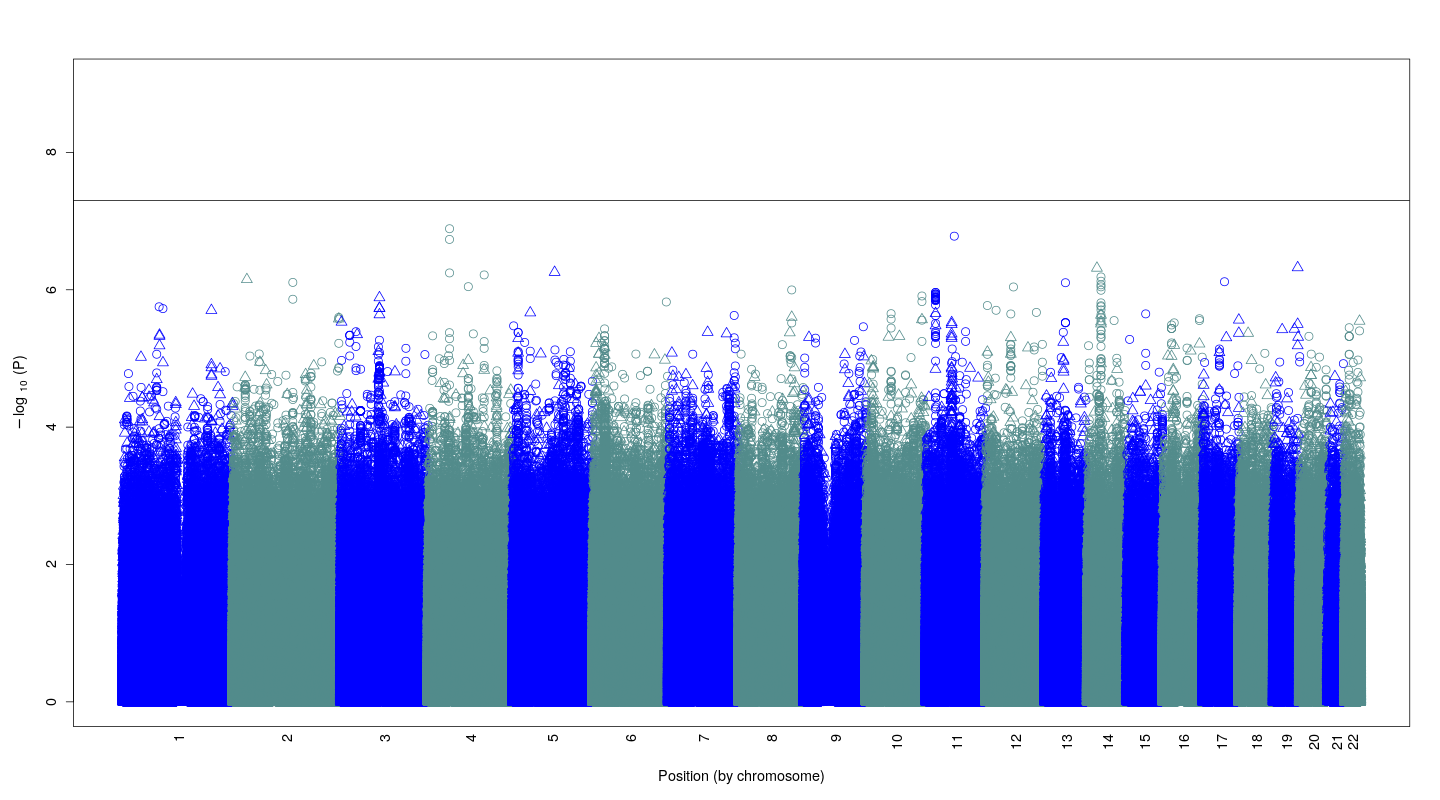

**Supplemental Figure 6.** Quantile-Quantile (QQ-plot) plot of GENOA African American + European American results.

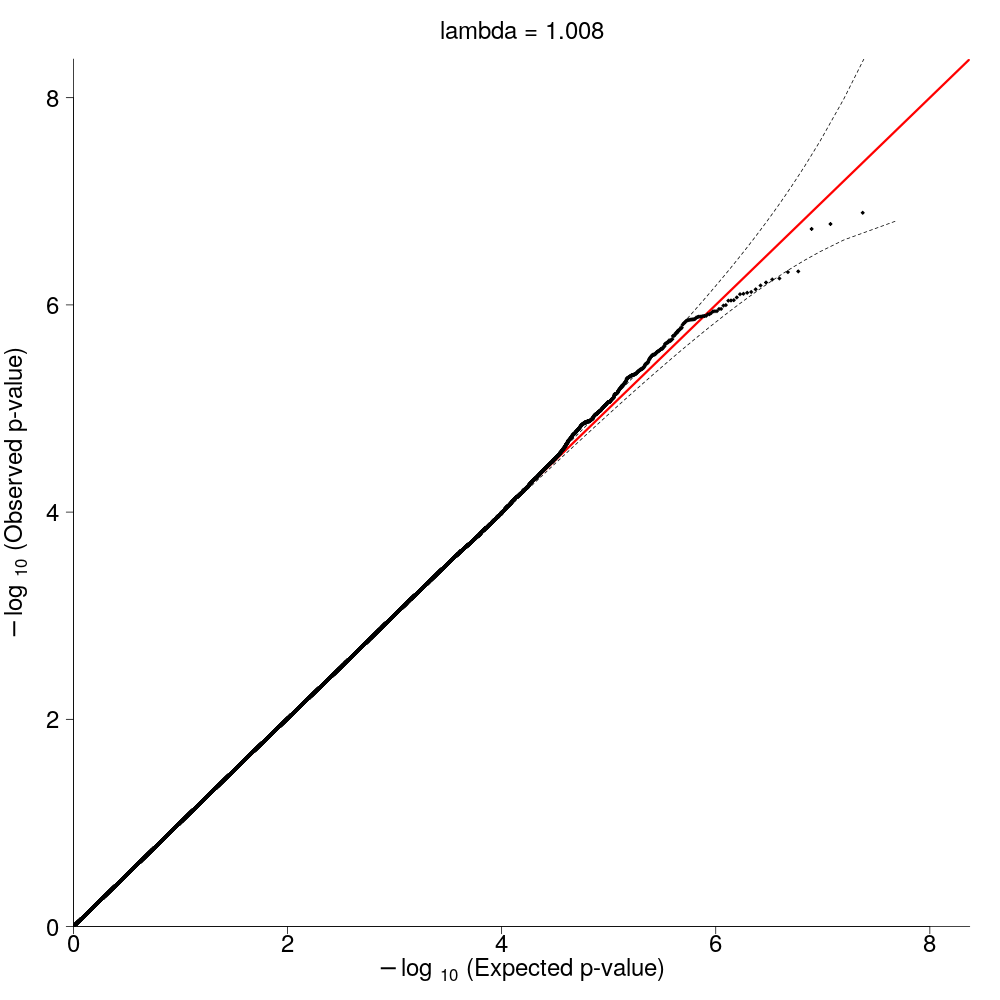

**Supplemental Figure 7.** LocusZoom plot of chromosome 4 genome-wide significant locus in GENOA European ancestry results.

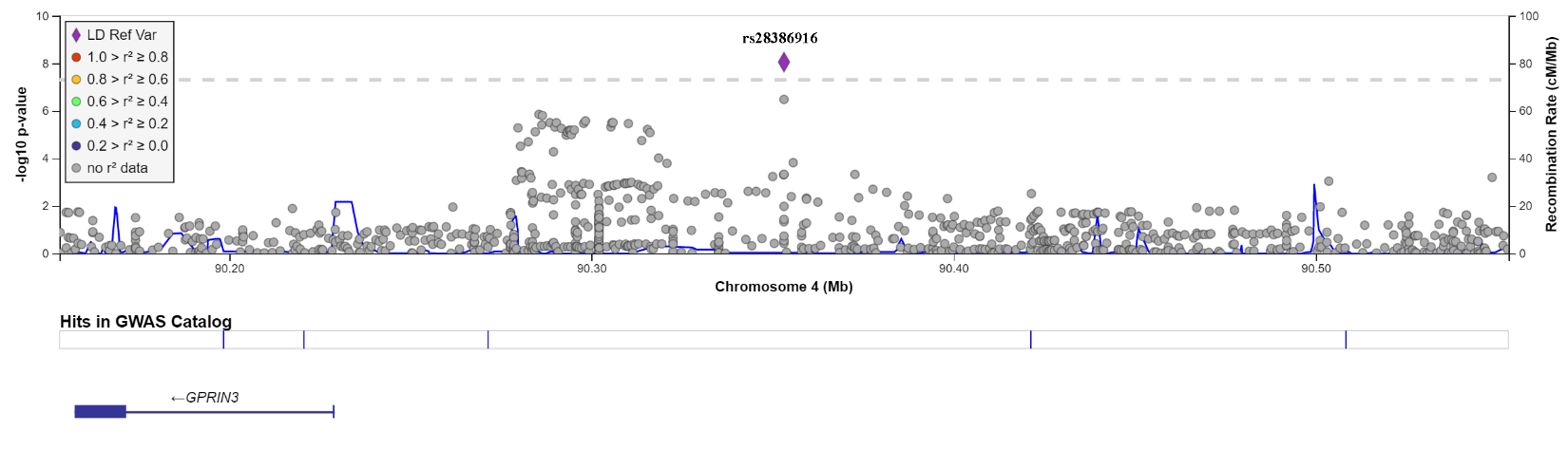

**Supplementary Figure 8.** Quantile-Quantile plot (QQ-plot) of the GenomicSEM results.

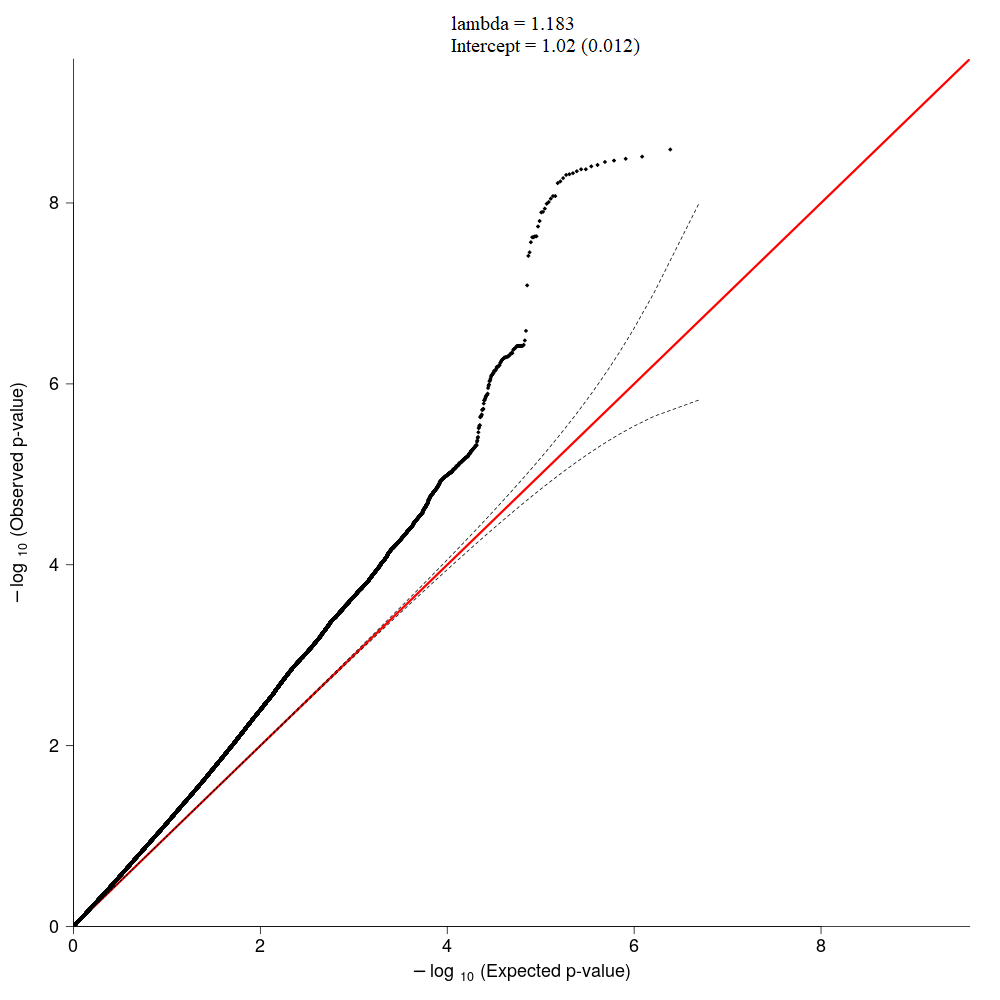

**Supplementary Figure 9.** Forest plot for **(A)** rs9478500 (genome-wide significant in our gSEM analysis) compared to the forest plot for **(B)** rs1799971. Sample sizes for the cohorts included in the GENOA meta-analysis (highlighted in blue) were Yale-Penn-CIDR (N=1,014), Yale-Penn-GO (N=895), VIDUS (N=2,175), UHS1 (N=9,245), ODB (N=2,765), Kreek (N=556), deCODE (N=275,468), COGA (n=7,631), CaTS2-MOLE (N=3,162), and CATS-PERTHUNT (N=1,920). Samples sizes for the cohorts included in the gSEM analysis were GENOA (N=304,831; effective N=28,428, PH (N=11,782; effective N=3,790), MVP12-YP-SAGE (N=82,707; effective N=35,799), PGC (N=8676; effective N=8,663), and finally the gSEM (N=403,915; effective N = 88,114) results.

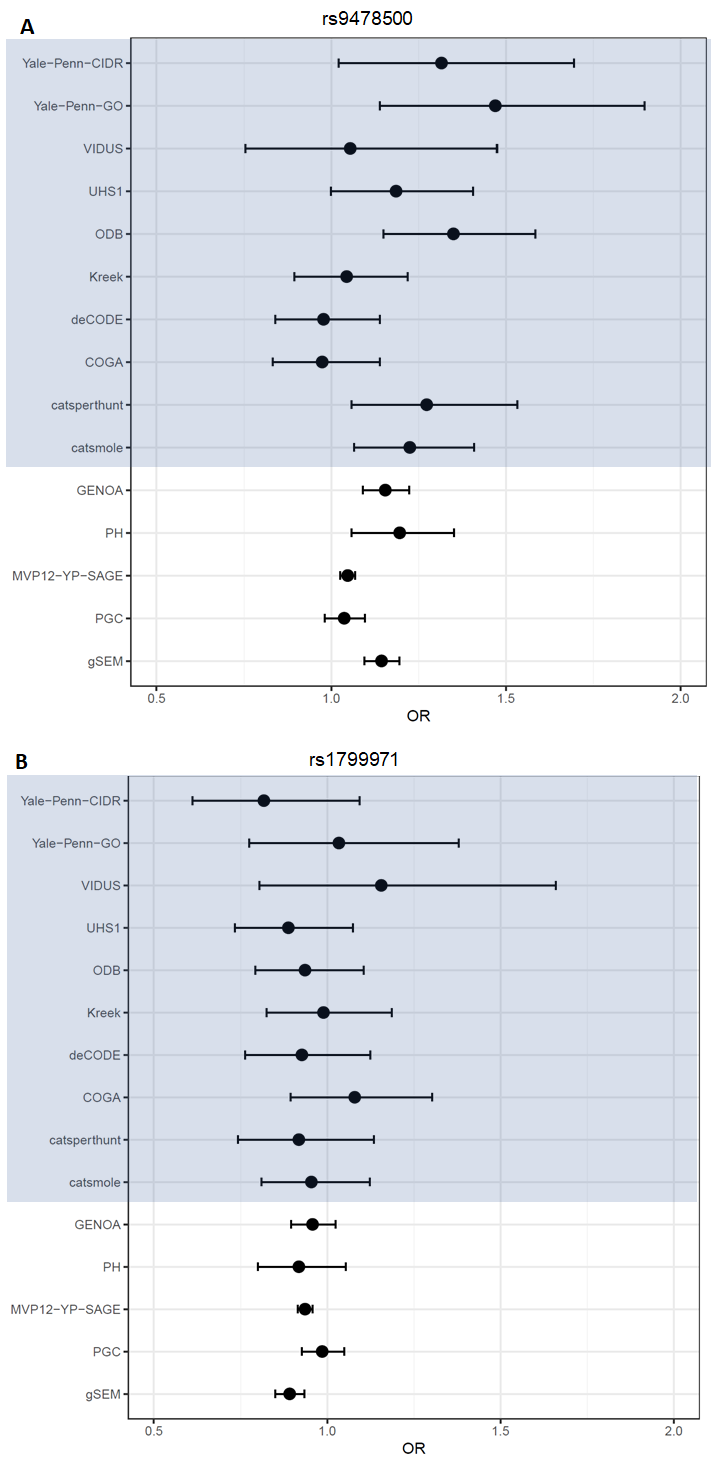

**Supplementary Figure 10.** Forest plot for genome-wide significant intergenic variant, rs13333582 (beta = -0.219, p=3.58x10^-8^) on chromosome 16. *This variant was not analyzed in the CaTS2-MOLE and CATS-PERTHUNT cohorts, therefore no data is available. Sample sizes for the cohorts included in the GENOA meta-analysis (highlighted in blue) were Yale-Penn-CIDR (N=1,014), Yale-Penn-GO (N=895), VIDUS (N=2,175), UHS1 (N=9,245), ODB (N=2,765), Kreek (N=556), deCODE (N=275,468), COGA (n=7,631), CaTS2-MOLE (N=3,162), and CATS-PERTHUNT (N=1,920). Samples sizes for the cohorts included in the gSEM analysis were GENOA (N=304,831; effective N=28,428, PH (N=11,782; effective N=3,790), MVP12-YP-SAGE (N=82,707; effective N=35,799), PGC (N=8676; effective N=8,663), and finally the gSEM (N=403,915; effective N = 88,114) results.

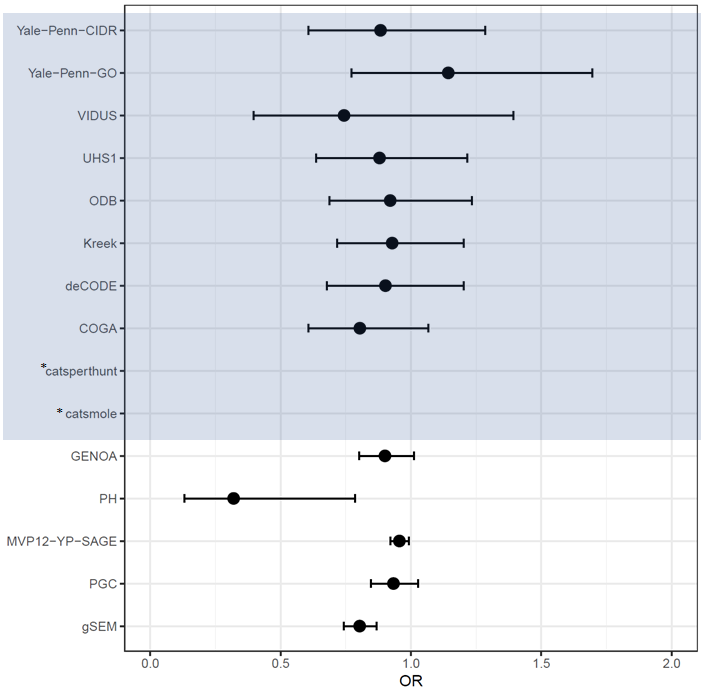

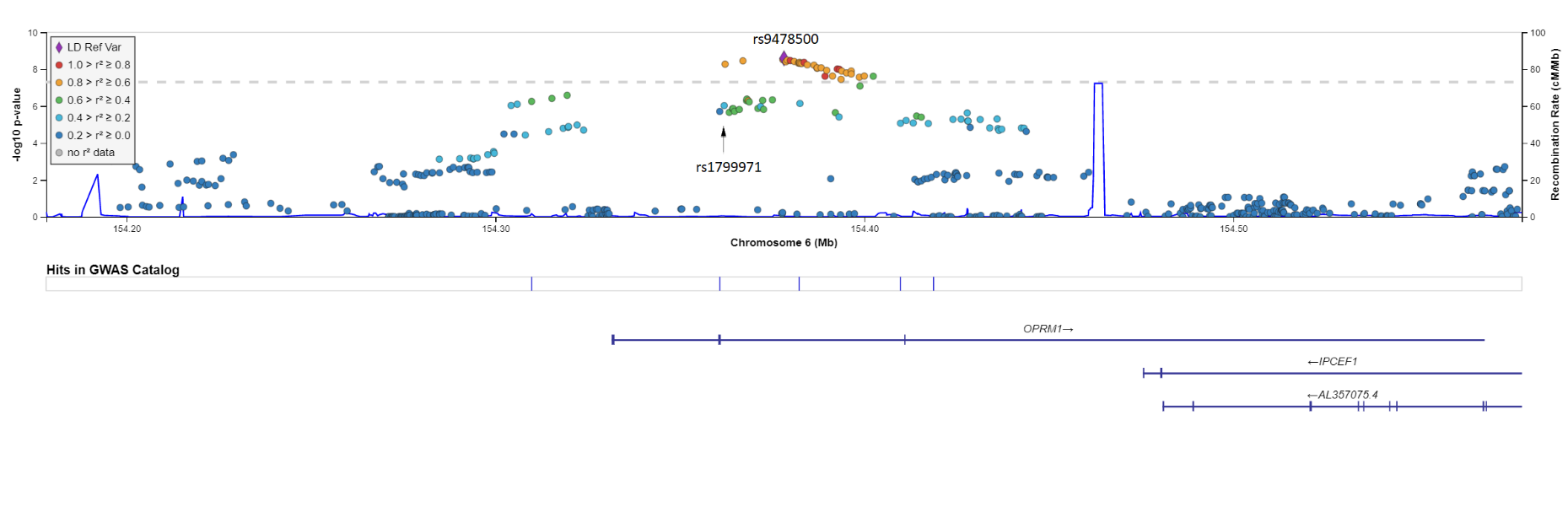
**Supplementary Figure 11.** LocusZoom plot of *OPRM1* region.

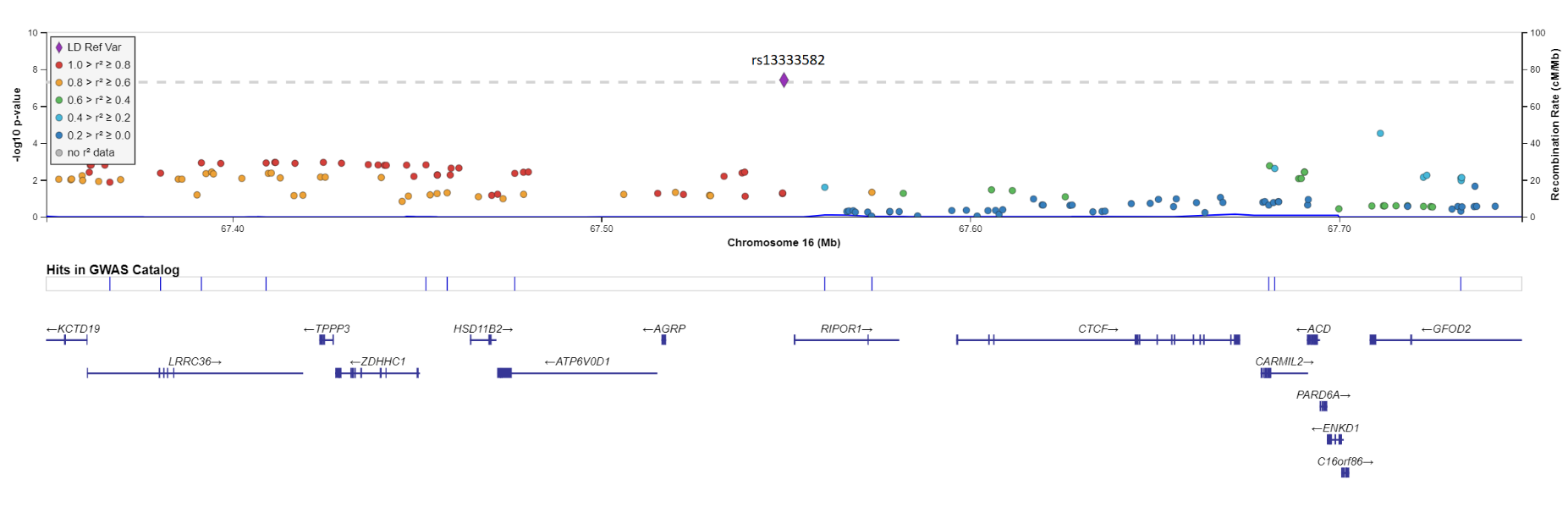
**Supplementary Figure 12.** LocusZoom plot of the genome-wide significant intergenic variant, rs13333582 (beta = -0.219, p=3.58x10^-8^) on chromosome 16.

**Supplementary Figure 13.** Quantile-quantile plot of the gene-based MAGMA analysis of the gSEM GWAS summary statistics. Note that the gSEM results used for this analysis show an elevated lambda (1.18) but an intercept near the null value of 1.0 (1.02), which indicated that the elevated lambda is due to association signal rather than bias. Thus, the elevated lambda in the MAGMA analysis is to be expected and unlikely to be due to bias.

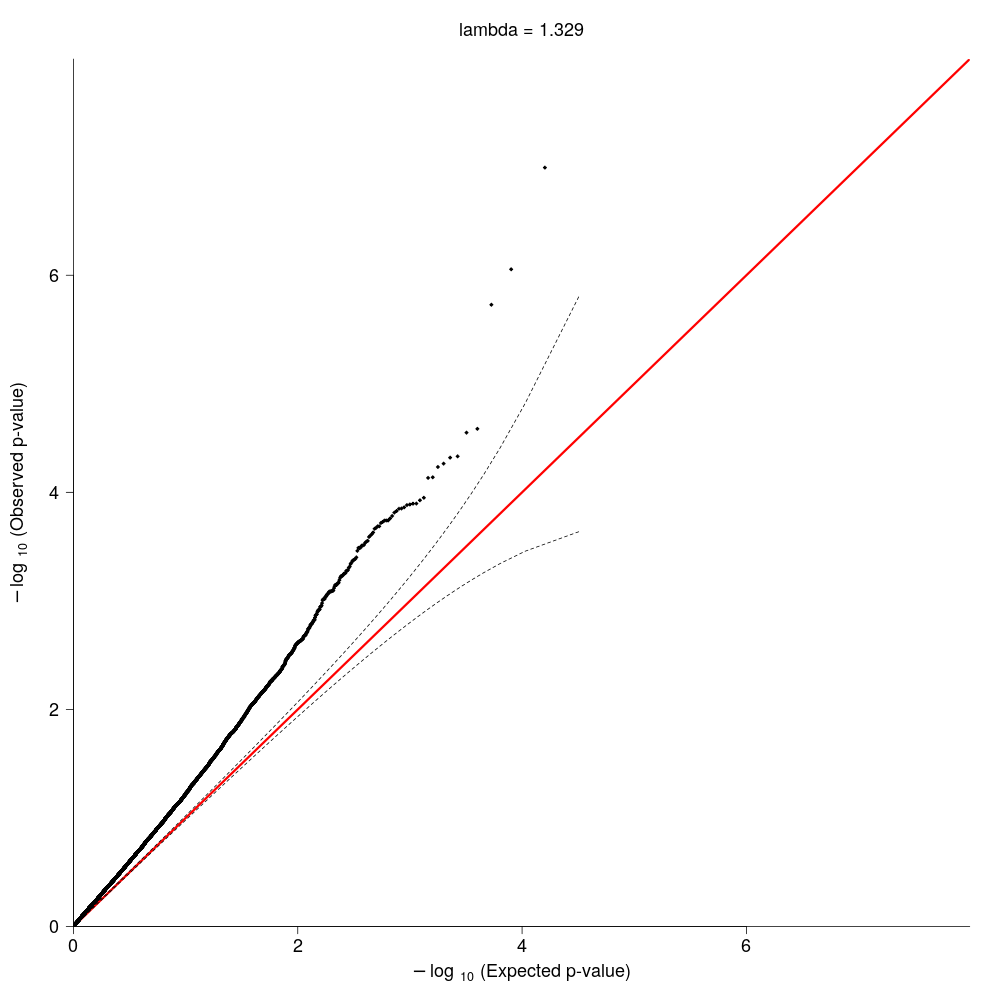

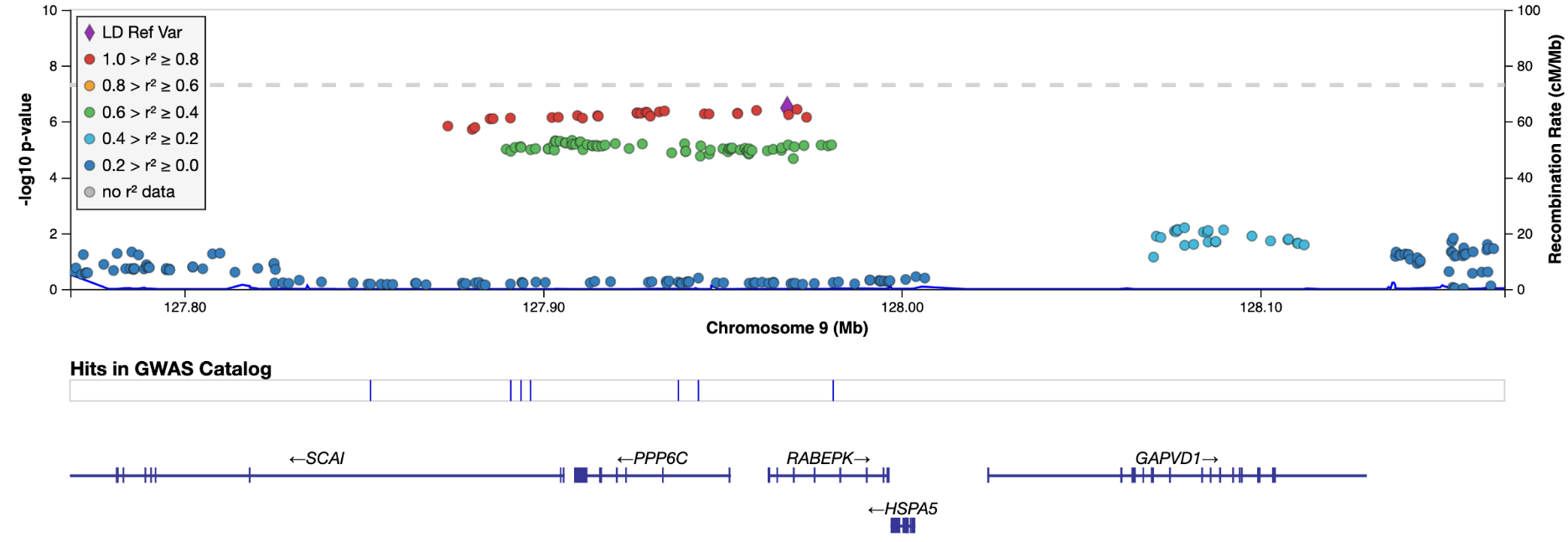
**Supplementary Figure 14.** LocusZoom plot for variants on chromosome 9 stretched across three genes, PPP6C, SCAI, and RABEPK.

**Supplementary Figure 15. LocusZoom plot for FURIN region for standard meta-analysis results.**

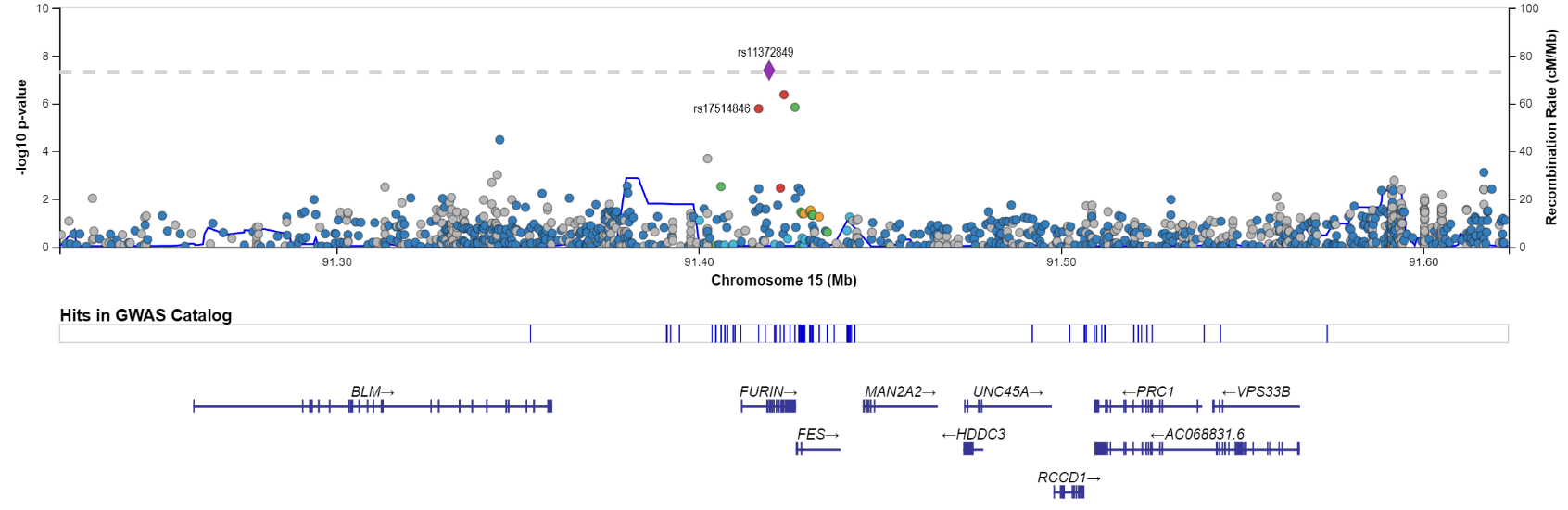

Note: rs17514846 is the variant included in the gSEM GWAS and drove the gene-based association in MAGMA.

**Supplementary Figure 16.** Forest plot for top variant, rs11372849, within *FURIN* on chromosome 15 from the standard meta-analysis results (beta=-0.074, p-value=4.11×10^-8^). *This variant was not analyzed in the PH cohort, therefore no data is available. Sample sizes for the cohorts included in this meta-analysis were Yale-Penn-CIDR (N=1,014), Yale-Penn-GO (N=895), VIDUS (N=2,175), UHS1 (N=9,245), ODB (N=2,765), Kreek (N=556), deCODE (N=275,468), COGA (n=7,631), CaTS2-MOLE (N=3,162), CATS-PERTHUNT (N=1,920), PH (11,782; effective N=2,790), MVP2, and MVP1.

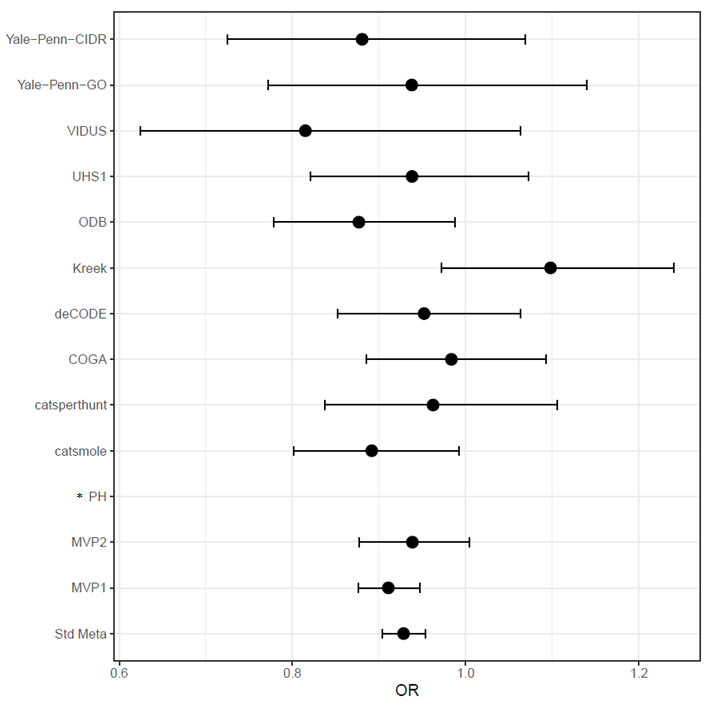

**Supplementary Figure 17.** Expression of (a) OPRM1, (b) SCAI, (c) PPP6C, (d) RABEPK, (e) FURIN Genes in GTEx Brain Tissues

A.

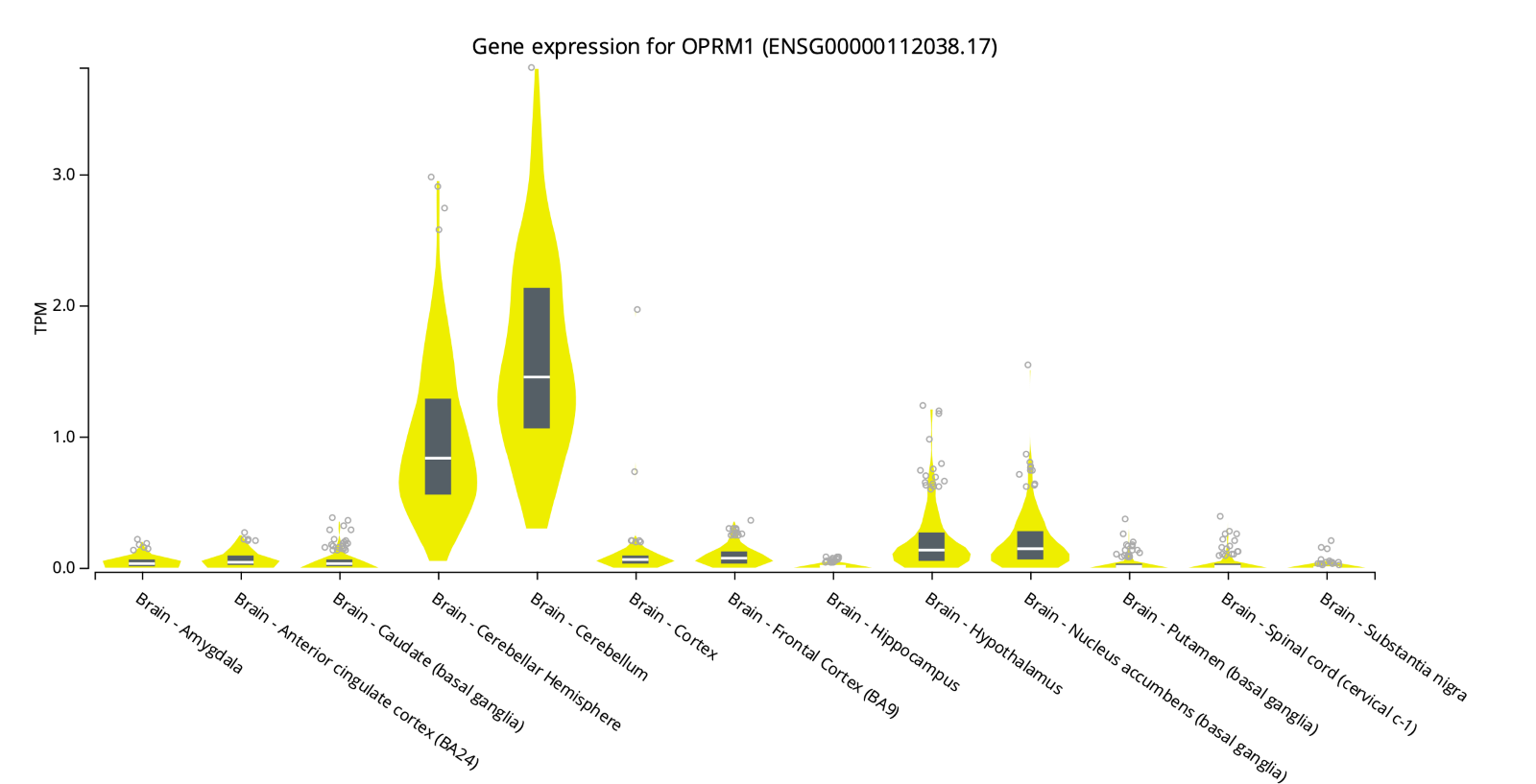

B.

 
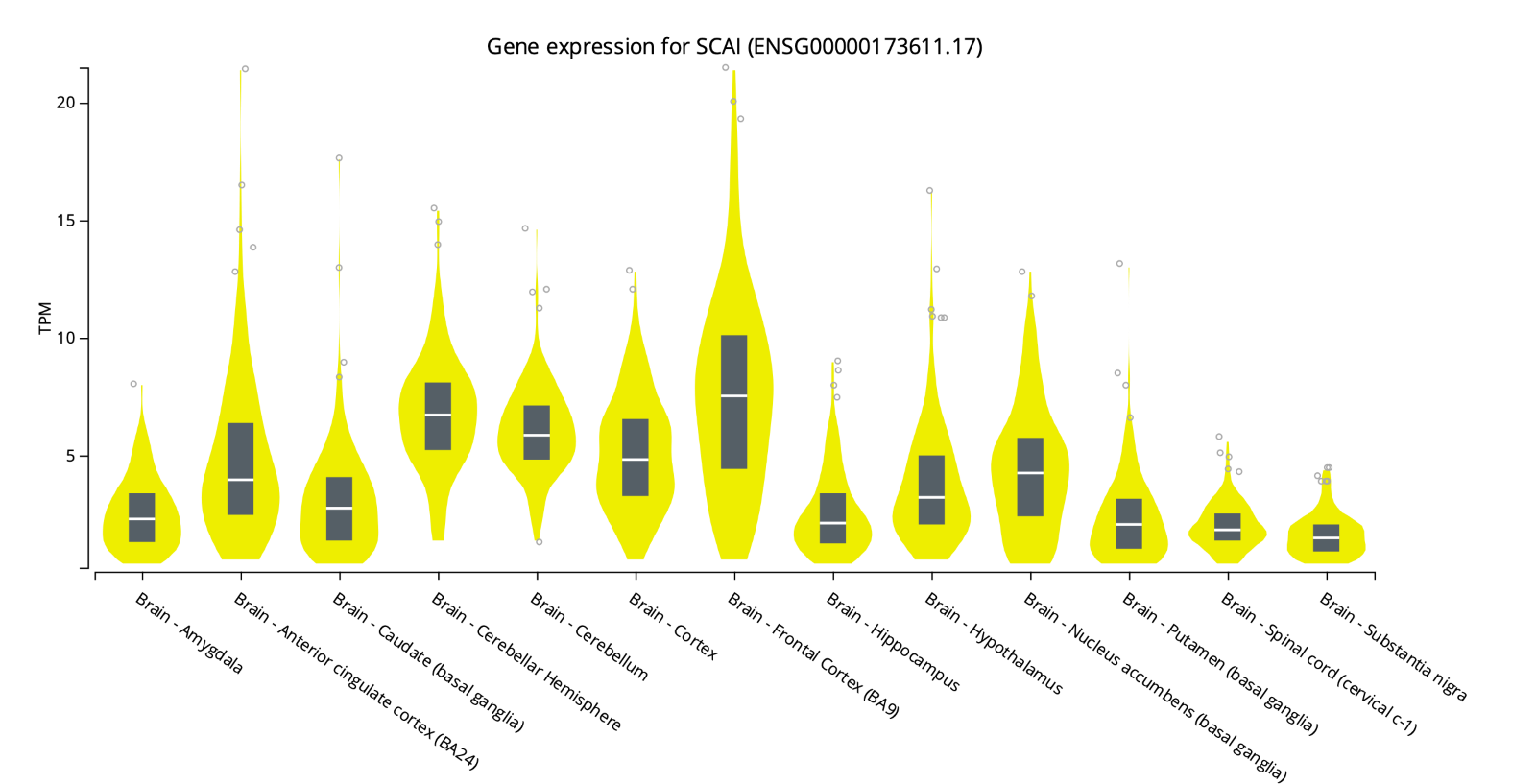

C.

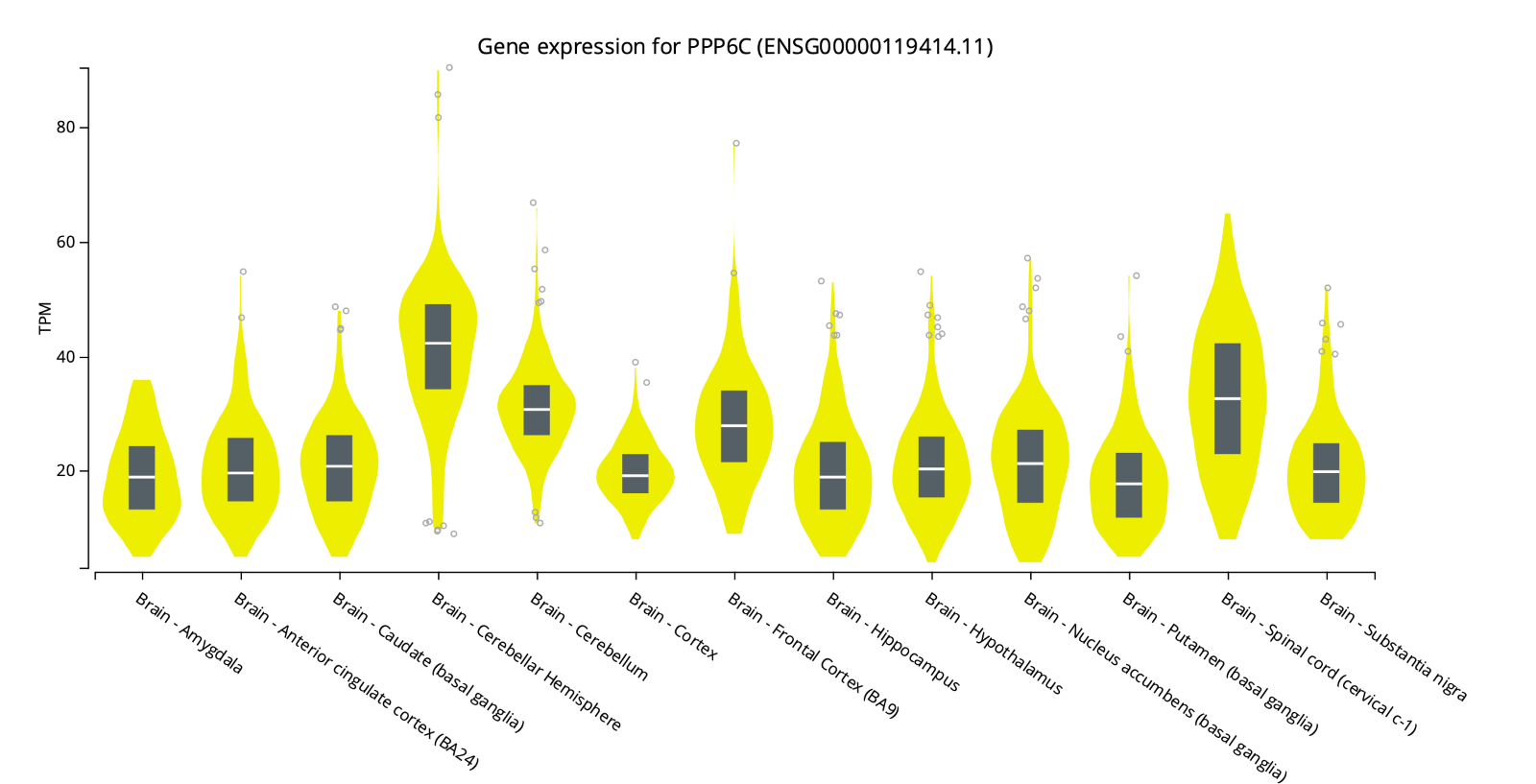

D.

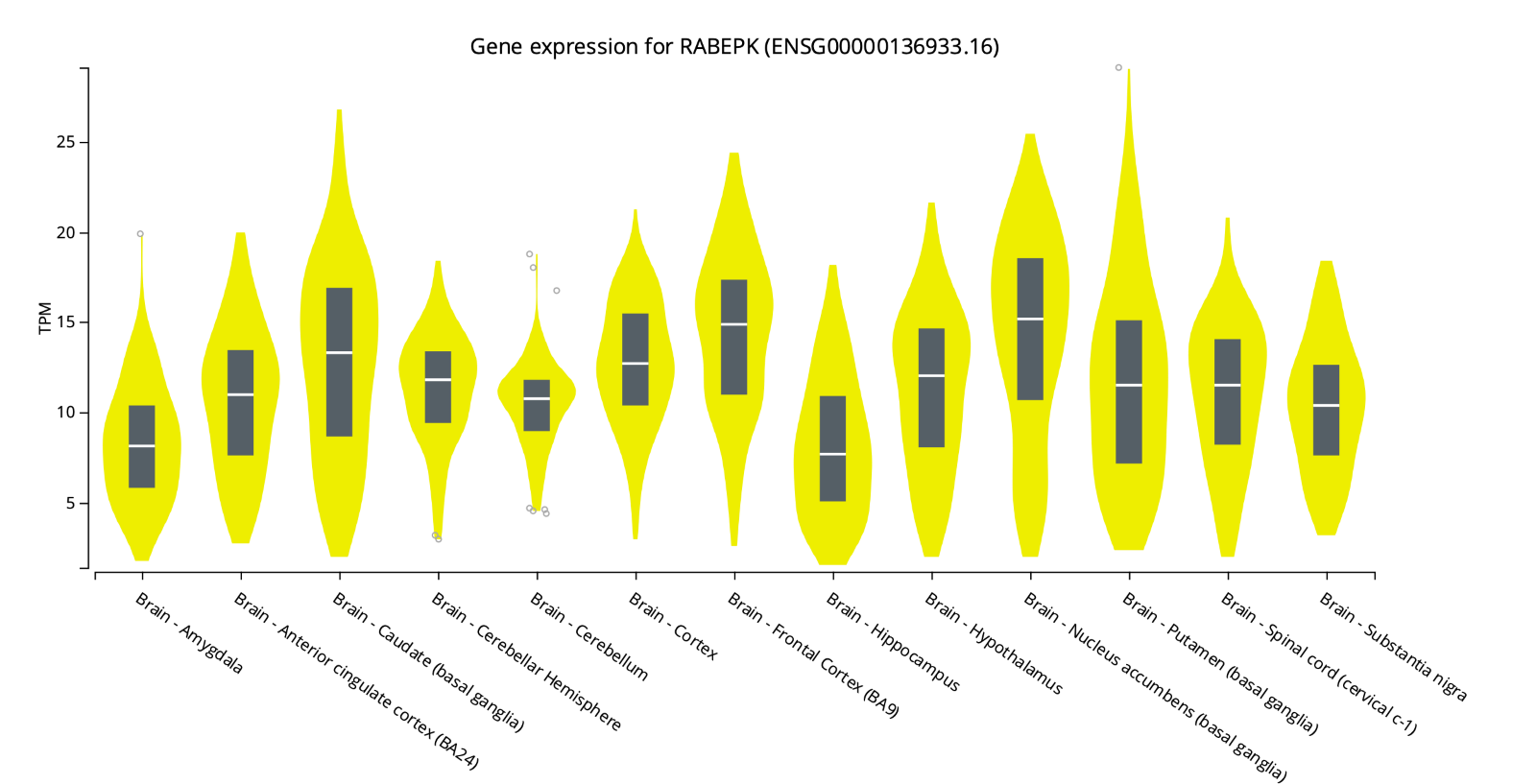

E.

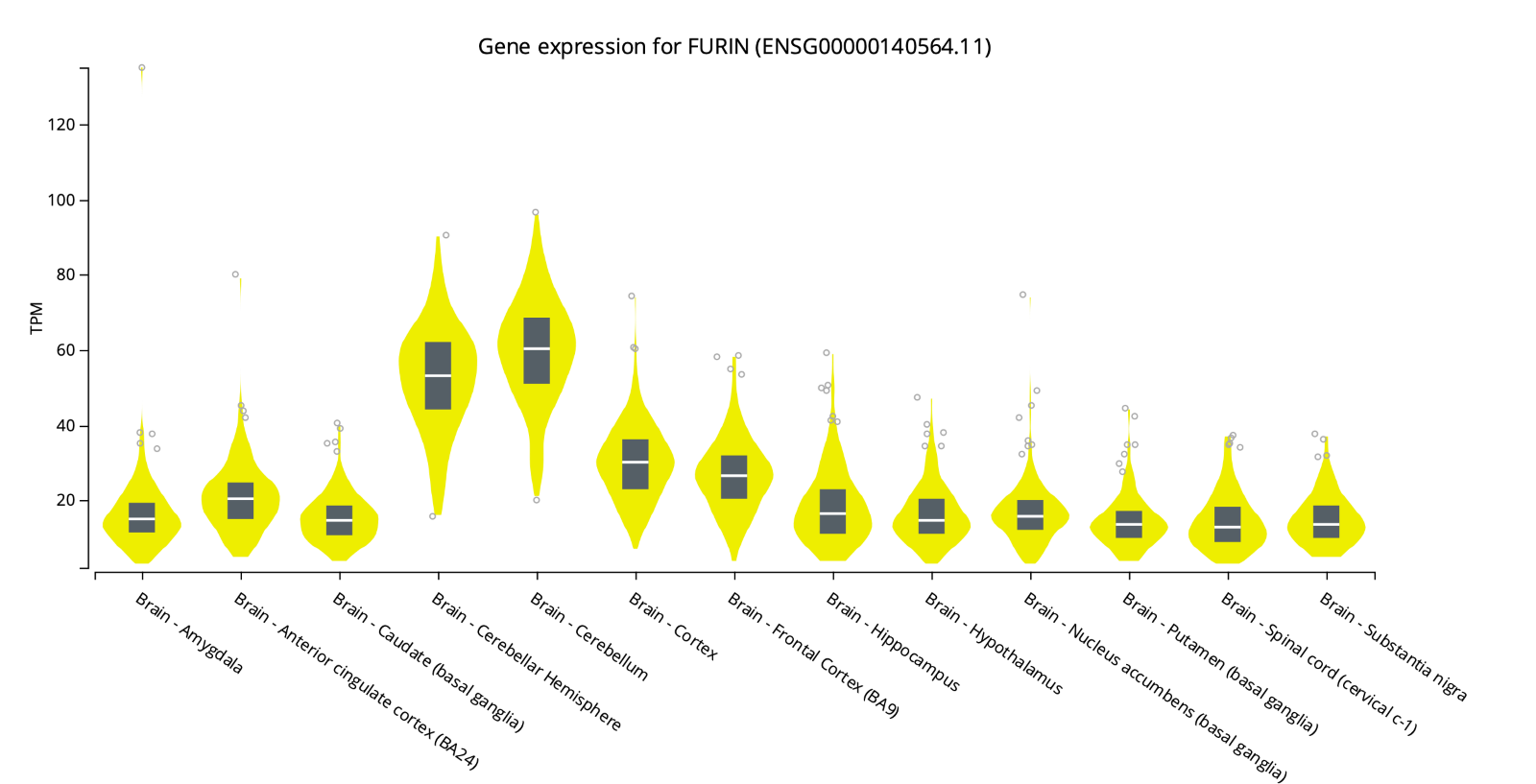

**Supplementary Figure 18. Open Target Pleiotropy plot for OPRM1**

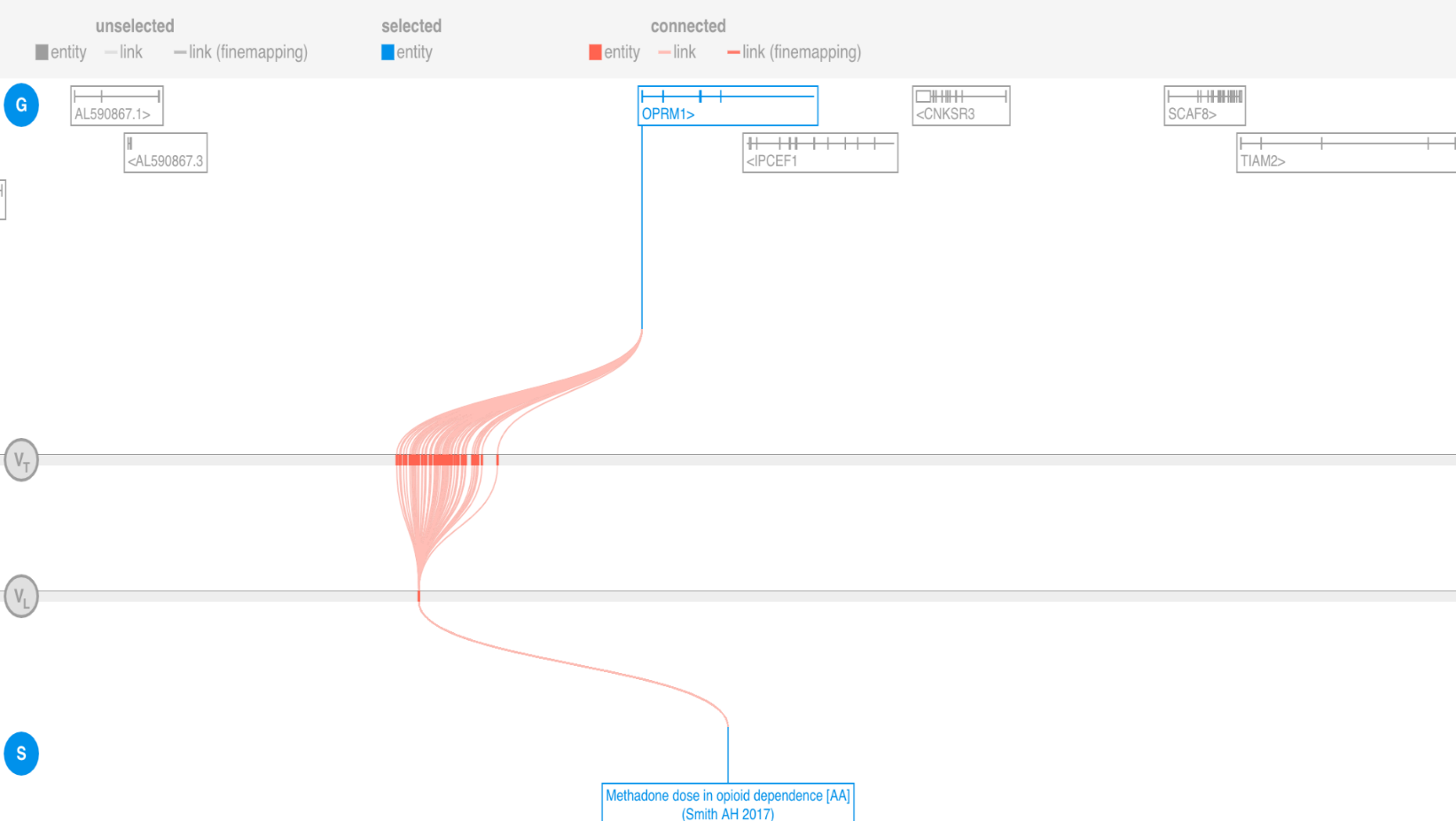

**Supplementary Figure 19. Open Targets Pleiotropy plot for PPP6C**

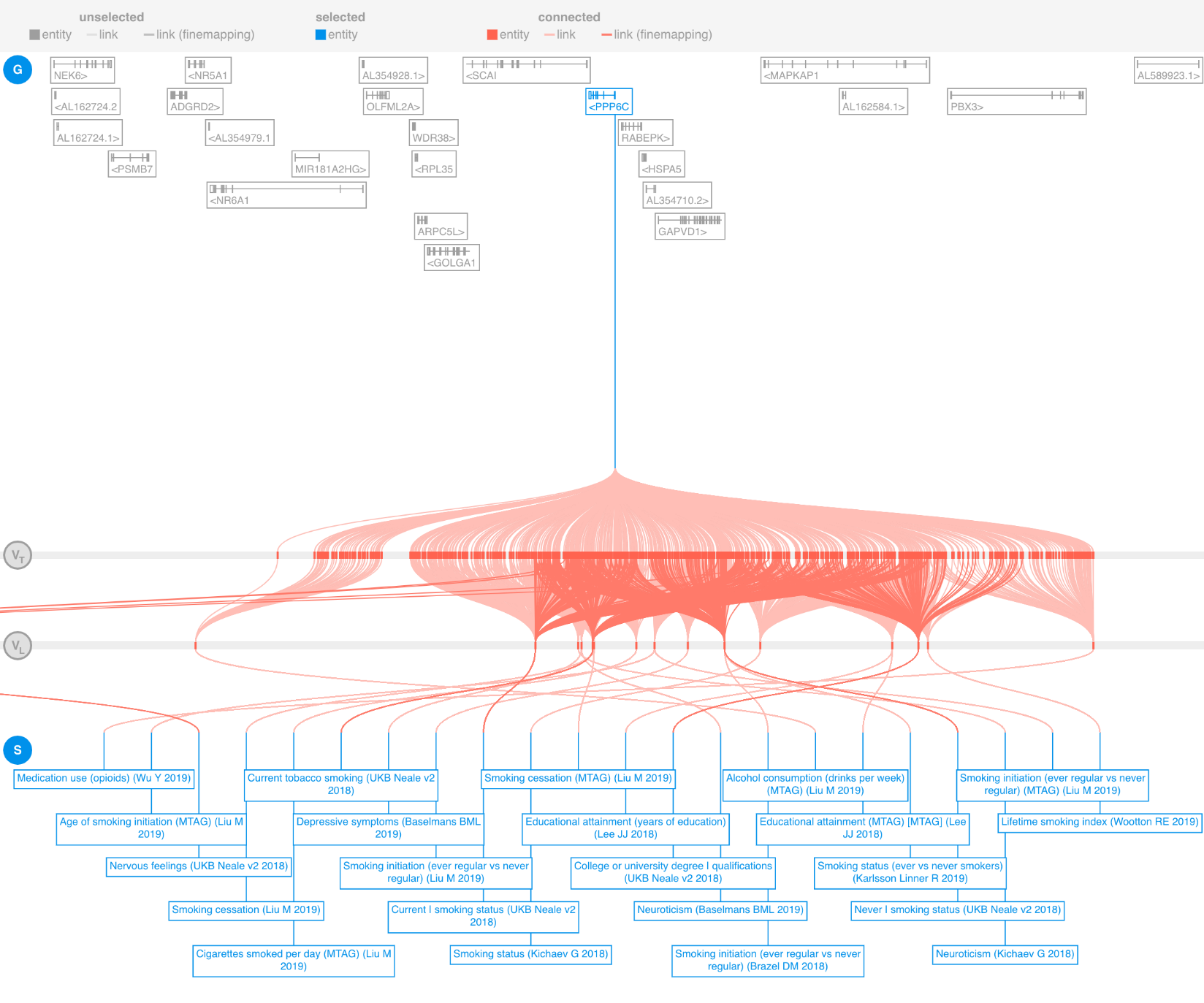

**Supplementary Figure 20.** Open Target pleiotropy plot for FURIN

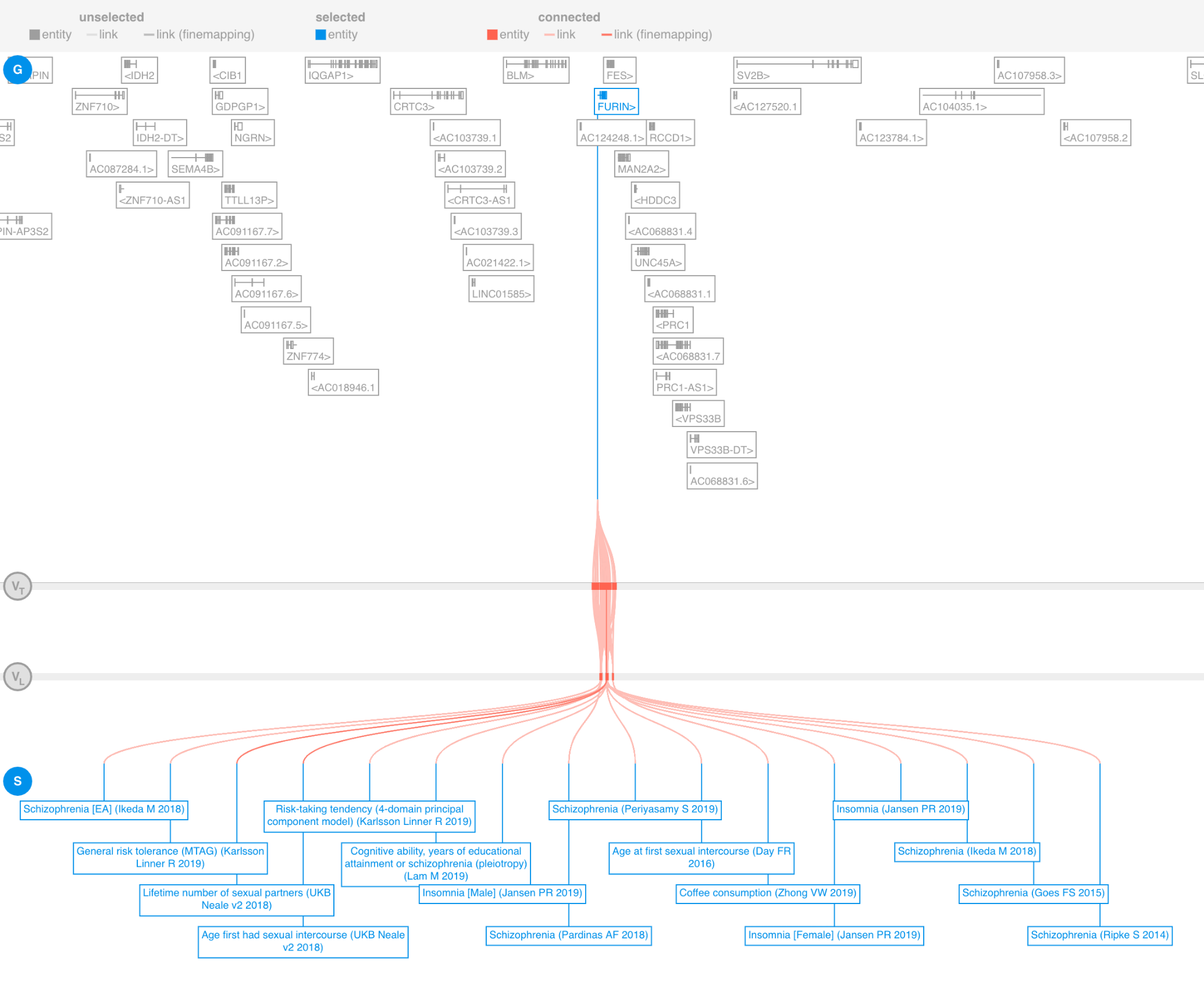
